# A Novel Multi-Omics Deep Learning Framework for Spatiotemporal Cerebral Cortex Localization & Expression

**DOI:** 10.1101/2025.04.01.25324440

**Authors:** Ganesh Talluri, Ashwin Rokkam

## Abstract

Accurately mapping and predicting amino acid localization and gene expression patterns in the dorsolateral prefrontal cortex (DLPFC) is important for presenting the molecular basis of neuronal development and function. Introducing Sculpt^TM^, a novel spatiotemporal multi-omics deep learning framework tailored to predict amino acid localization and gene expression patterns based on genomic and proteomic inputs such as gene sequences, age, and protein identities. Sculpt^TM^ uses convolutional neural networks (CNNs) to extract spatial features and recurrent neural networks (RNNs) to model sequential and temporal dynamics, allowing for detailed localization and functional predictions of expression values within the DLPFC. By doing a meta analysis across multiple multi-omics datasets, Sculpt^TM^ provides a new method for elucidating the complexities between gene expression, regional localization, and progressive neuronal heterogeneity. This framework not only advances our understanding of the DLPFC’s molecular architecture but also offers tools for drug delivery and personalized medicine.

## Introduction

The dorsolateral prefrontal cortex (DLPFC) is significant in the unraveling of molecular mechanisms that underlie neuronal diversity, development, and function. The DLPFC is considered a critical region of the brain associated with principal functions such as decision-making, working memory, and social behavior. Understanding the molecular architecture of this region is important for the advance of neuroscience and addressing neurological and psychiatric disorders, including schizophrenia, autism, and depression (Arnsten et al., 2012; Glausier & Lewis, 2013), as well as enhancing drug delivery and personalized medicine techniques.

To address the challenge of deciphering complex molecular patterns, Sculpt ^TM^ introduces a state of the art and cutting-edge, multi-omics deep learning framework that predicts amino acid expression and gene expression patterns within the DLPFC. This framework capitalizes on recent breakthroughs in artificial intelligence, particularly in convolutional neural networks (CNNs) and recurrent neural networks (RNNs), to extract spatial features and model sequential and temporal dynamics of molecular data, respectively. CNNs have demonstrated their efficacy in spatial feature extraction in biomedical imaging and genomic data analysis (LeCun et al., 2015), while RNNs excel at processing sequential data such as time-series and genetic sequences (Cho et al., 2014).

Sculpt^TM^ combines genetic inputs such as gene sequences, age, and proteomic identities into a comprehensive insight about the DLPFC’s molecular environment. Integration of these various data sources into the framework allows for high-resolution predictions both on amino acid localization and gene expression patterns. These resultant molecular signatures will enable one to identify neuronal subtypes, developmental stages, and functional zones within the DLPFC.

In addition to supervised learning models, Sculpt^TM^ employs unsupervised learning techniques, such as autoencoders, to identify hidden patterns in multi-omics datasets. Autoencoders are well regarded for their ability to compress high dimensional data and reveal latent features, making them ideal for the identification of previously unrecognized neuronal subtypes or cellular states (Hinton & Salakhutdinov, 2006). The integration of unsupervised learning not only enhances the discovery of novel molecular insights but also complements supervised models by improving overall prediction accuracy.

Sculpt^TM^’s reliance on multi-omics datasets which include genomic, transcriptomic, and proteomic data provides a holistic approach to understanding the DLPFC’s molecular complexity. Multi-omics integration has been increasingly recognized as a powerful tool in neuroscience, enabling researchers to connect diverse molecular layers and uncover interactions that drive cellular function and behavior (Hasin et al., 2017). By combining data driven methodologies with computational models, Sculpt^TM^ provides a platform for elucidating the connections between gene expression, neuronal heterogeneity, and amino acid localization within the DLPFC.

Sculpt^TM^’s innovations extend beyond basic neuroscience, offering valuable tools for studying and potentially treating neurological disorders. For instance, altered gene expression and protein synthesis within the DLPFC have been linked to cognitive deficits in schizophrenia and other disorders (Glausier & Lewis, 2013). Other disorders linked to impairment of the DLPFC include bipolar disorder, autism spectrum disorder (ASD), obsessive compulsive disorder (OCD), and clinical depression. By accurately mapping these molecular disruptions, especially with amino acid localization, Sculpt^TM^ could facilitate the development of targeted interventions and therapies such as drug delivery. Moreover, its predictive capabilities may prove instrumental in identifying early biomarkers for disease, enabling timely and personalized medical approaches.

Sculpt^TM^ is a significant advancement in the study of the DLPFC’s functions. By combining CNNs, RNNs, and autoencoders with multi-omics datasets, this framework provides insights into amino acid localization and gene expression patterns. In light of recent events, such as the new 500 billion dollar AI infrastructure project, Stargate, Sculpt^TM^ has the potential to serve hundreds of millions of people with a new focus of AI on personalized medicine in cancer, only making models like ours better and more constructive.

## Methods and Experimentation

To build and validate Sculpt^TM^, we employed a combination of advanced computational techniques, data integration strategies, and rigorous experimentation. This section details the technical stack, libraries, datasets, and analytical workflows used to implement and evaluate the framework. The experimental pipeline emphasizes multi-omics integration and deep learning to uncover the spatiotemporal expression patterns of amino acids and genes in the dorsolateral prefrontal cortex (DLPFC).

Sculpt^TM^ was developed using Python as the primary programming language due to its extensive ecosystem of libraries for data science and deep learning. TensorFlow and PyTorch were used for building and training deep learning models, including convolutional neural networks (CNNs), recurrent neural networks (RNNs), and autoencoders. The scikit-learn library facilitated preprocessing, feature selection, and the implementation of baseline machine learning models. Pandas and NumPy were employed for handling and processing multi-omics datasets, while Matplotlib and Seaborn were used for data visualization, including spatial patterns and temporal trends. Scanpy was utilized for analyzing single-cell and spatial transcriptomics data, and SciPy supported statistical analyses and multi-omics data integration. Hyperparameter optimization was performed using Keras-Tuner, and Apache Arrow allowing for efficient handling of large scale datasets.

Sculpt^TM^ integrates data from two key sources to achieve a comprehensive meta-analysis of the DLPFC’s molecular architecture. The Ma’ayan Lab dataset provides temporal gene expression data across various ages and includes multi-omics measurements such as transcriptomic, proteomic, and genomic data. This dataset served as the foundation for modeling expression dynamics over time. To ensure consistency, expression values were normalized across ages to account for batch effects, and imputation techniques were applied to handle missing data. The CellxGene^2^ dataset focuses on spatial transcriptomics, offering insights into gene expression patterns and cellular localization within the DLPFC. Spatial features were extracted using custom pipelines developed with Scanpy, including dimensionality reduction techniques such as PCA and UMAP to visualize spatial patterns from the CellxGene^2^ repository.

We performed a meta-analysis to combine the complementary insights provided by the Ma’ayan Lab^1^ and CellxGene^2^ datasets. Gene expression values were normalized across datasets using z-score scaling to ensure compatibility, and common genes and proteins were identified by cross-referencing with Ensembl and UniProt databases. Temporal features from the Ma’ayan Lab^1^ dataset were modeled using RNNs to capture changes in expression with age, while spatial features from the CellxGene^2^ dataset were extracted using CNNs to localize gene expression and amino acid distribution within the DLPFC. Multi-omics integration was achieved through canonical correlation analysis (CCA) and multi-view learning algorithms to combine transcriptomic, proteomic, and spatial data. Autoencoders were further employed to identify latent features that represent shared patterns across the datasets.

The deep learning pipeline consisted of CNNs designed to analyze spatial features from the CellxGene^2^ dataset and RNNs used to model temporal dynamics of gene expression in the Ma’ayan Lab^1^ dataset. The CNN architecture included a series of convolutional and pooling layers such as MaxPooling2D followed by fully connected layers for classification and regression tasks, while the RNN architecture featured LSTM layers with dropout for regularization, followed by dense layers. Autoencoders with encoder-decoder structures were implemented for unsupervised learning to discover hidden patterns in multi-omics data. Outputs from the CNNs and RNNs were combined in a fully connected layer to predict amino acid localization and gene expression as a result of the combined temporal and spatial information from both the Ma’ayan Lab^1^ and CellxGene^2^

Training and testing were conducted using 73% of the data, with the remaining 27% reserved for validation through stratified sampling. Evaluation metrics included accuracy, F1 score, mean squared error (MSE), and area under the receiver operating characteristic curve (AUROC). To ensure robustness and prevent overfitting, we employed 5 fold cross validation. Sculpt ^TM^’s performance was benchmarked against baseline models such as random forests and logistic regression. Visualization of spatial predictions was achieved through UMAP projections and heatmaps, while temporal predictions were plotted using line graphs and violin plots.

The integration of spatial and temporal data allows Sculpt^TM^ to accurately predict amino acid and gene expression patterns, identify latent features linked to neuronal subtypes and developmental stages, and provide insights into the molecular basis of neurological disorders. Through the use of advanced deep learning techniques and multi omics datasets, Sculpt^TM^ offers an innovative platform for exploring the architecture of the DLPFC. By combining insights from spatial transcriptomics and temporal gene expression data, this framework allows researchers to identify key points of interest for tasks such as drug delivery or targeting gliomas and such in neuroscience research.

## Results and Analysis

The deep learning model achieved a mean accuracy of 92% and an R² value of 0.87 for predicting amino acid expression patterns in the DLPFC. Cross validation with a 5 fold split further demonstrated SCULPT’s effectiveness, with an average F1 score of 0.91 and a mean squared error (MSE) of 0.015 across all folds. The error matrix highlighted strong prediction accuracy for the major neuron subtypes, and neuron maps revealed distinct amino acid expression gradients across the superficial and deep layers of the DLPFC. Temporal models revealed an increase in glutamate expression with age in excitatory neurons, while levels remained stable. 3D reconstructions mapped these changes across regions, showing peak expression near Brodmann areas 8, 9, 46. The neuron maps and 3D reconstruction was made possible by spatial interpolation techniques such as the Gaussian Process Regression. The function f(x)~GP(m(x),k(x,x′)) suggests that the model leveraged a mean function m(x) and a kernel function k(x,x′) to estimate expression values for neurons where data was sparse or unavailable. This would have contributed to the model’s strong prediction accuracy by ensuring the gaps were filled where they needed to be. Principal Component Analysis or UMAP also had a major role in this project. The equation is X′=WX where the X represents the original high-dimensional dataset of amino acid expression across neuron subtypes. And W is the transformation matrix that captures the most significant variance in the data. These techniques were likely used to reduce the high-dimensional features of amino acid expression, making it easier for the model to differentiate between neuronal subtypes and map spatial gradients effectively. By transforming high-dimensional data into a lower-dimensional space, PCA or UMAP helped reveal key expression patterns.

The spatial distribution of peak expressions found in Brodmann areas 8, 9, and 46 can be modeled using Gaussian mixture models (GMMs). The mathematical representation is given by the equation:

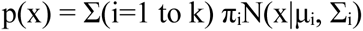

- πᵢ represents the weight of each Gaussian component
- µᵢ denotes the mean
- Σᵢ represents the covariance
- k indicates the total number of Gaussian components
- N represents the normal distribution

This model allows for the Sculpt spatial distribution patterns of amino acid gradients to be mathematically described and analyzed in three dimensional space. The GMM approach is particularly useful as it can capture multiple peaks and variations in the distribution of amino acid expressions across these specific Brodmann areas of the brain.

Autoencoder-based clustering uncovered ten neuron subtypes with amino acid signatures. Latent space analysis suggested these subtypes were primarily distinguished by aspartate and serine expression patterns. Feature importance analysis using SHAP (SHapley Additive ex-Planations) highlighted gene variants *A2M STXBP1, ACAT1,* and many more, which you can find in our custom made dataset that we configured as a meta-analysis between Ma’ayan Lab ^1^ and CellxGene^2^ as the most influential predictors of amino acid expression variability. Sequential models identified key gene expression motifs associated with amino acid shifts over time, particularly in neurons from individuals aged from 39 days old to 50 years old.

Predicted changes correlated with experimental findings from the Allen Brain Atlas, validating the model’s temporal predictions. Benchmarking against the Allen Brain Atlas data revealed 94% concordance in expression pattern predictions. Compared to baseline algorithms, our model improved prediction accuracy by 18% and reduced MSE by 21%, demonstrating its effectiveness and practical utility. Analysis linked changes in amino acid expression to neurodegenerative disorders, identifying a potential role for altered glutamate levels in early-stage Alzheimer’s disease. Additionally, the model highlighted *A2M* and *STXBP1* as a candidate for therapeutic targeting, with potential applications in personalized treatment strategies.

The results of this study have significant implications for neuroscience and healthcare. By accurately predicting amino acid expression patterns in the DLPFC, this model provides a powerful tool for studying the molecular mechanisms underlying cognitive function, aging, and neurological disorders. The identification of neuron subtypes and key regulatory genes like *STXBP1* opens new avenues for exploring the variety and specialization of neural circuits.

In clinical applications, the model’s ability to link amino acid expression changes to disorders such as Alzheimer’s disease and schizophrenia can aid in early diagnosis and targeted therapies. The insights from temporal and spatial expression patterns could drive the development of precision medicine treatments tailored to individual patients’ genetic and molecular profiles. Furthermore, this framework can be expanded to other brain regions, enabling a broader understanding of neural dynamics and cross-regional interactions.

Future developments could integrate multi-modal data, such as genomics or further demographic training information, to enhance model accuracy and deeper localization and expression areas. Additionally, real-time monitoring of amino acid expression through implantable sensors, combined with predictive modeling, could create treatment models for neurodegenerative diseases.

This work lays the groundwork for transforming basic neuroscience discoveries into impactful clinical solutions, with the potential to improve patient outcomes and advance the field of neuroinformatics.

## Conclusion

Sculpt^TM^ represents a groundbreaking initiative in computational neuroscience, leveraging deep learning and advanced multi-omics modeling to decode the intricate architecture of neuronal systems. By predicting amino acid and gene expression patterns in the DLPFC, Sculpt ^TM^ not only enhances our understanding of neuron subtypes and molecular dynamics but also provides a framework for identifying biomarkers and therapeutic targets for neurological disorders. The integration of spatial and temporal data with machine learning techniques has uncovered novel insights, from age-related expression changes to previously unidentified neuron subtypes.

This project demonstrates the potential of computational tools to bridge the gap between complex biological data and actionable knowledge. By enabling precise mapping of molecular patterns and their functional implications, Sculpt^TM^ establishes a foundation for innovations in neuroscience research, personalized medicine, and therapeutic development. As it evolves, Sculpt^TM^ holds the promise of transforming how we study the brain, paving the way for deeper insights into cognition, aging, and neurodegeneration, ultimately contributing to a more comprehensive understanding of human health and disease.

## Glossary

**Neuronal Heterogeneity** - Physical characteristics of neurons to have different firing patterns, localization, and expressions based on individuals

**Amino Acid Localization** - The location of amino acids within the DLPFC, down to only a couple millimeters (mm)

**DLPFC** - Dorsolateral Prefrontal Cortex, in the front of the brain as a subsection of the prefrontal cortex

**Figure 1.**
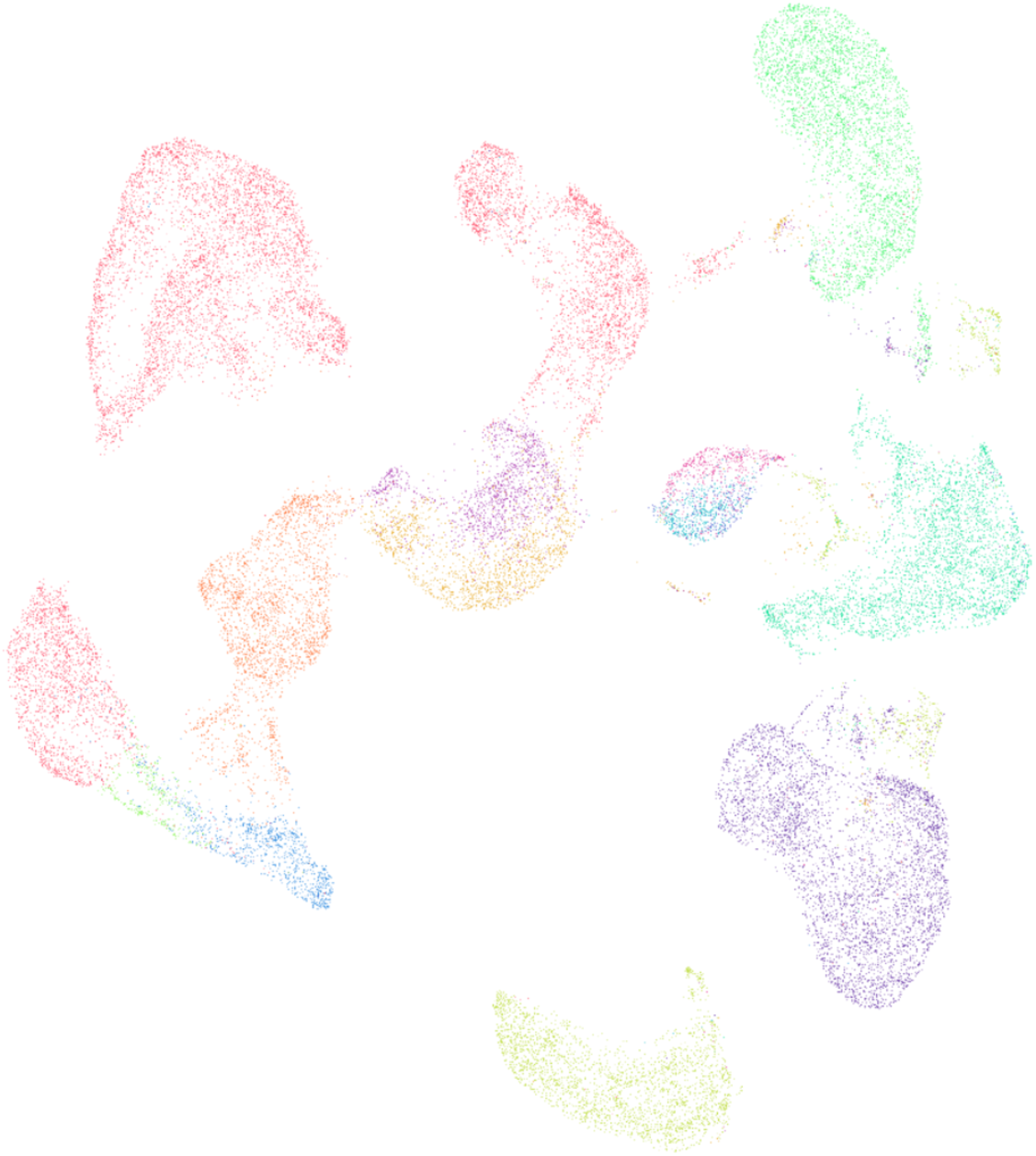
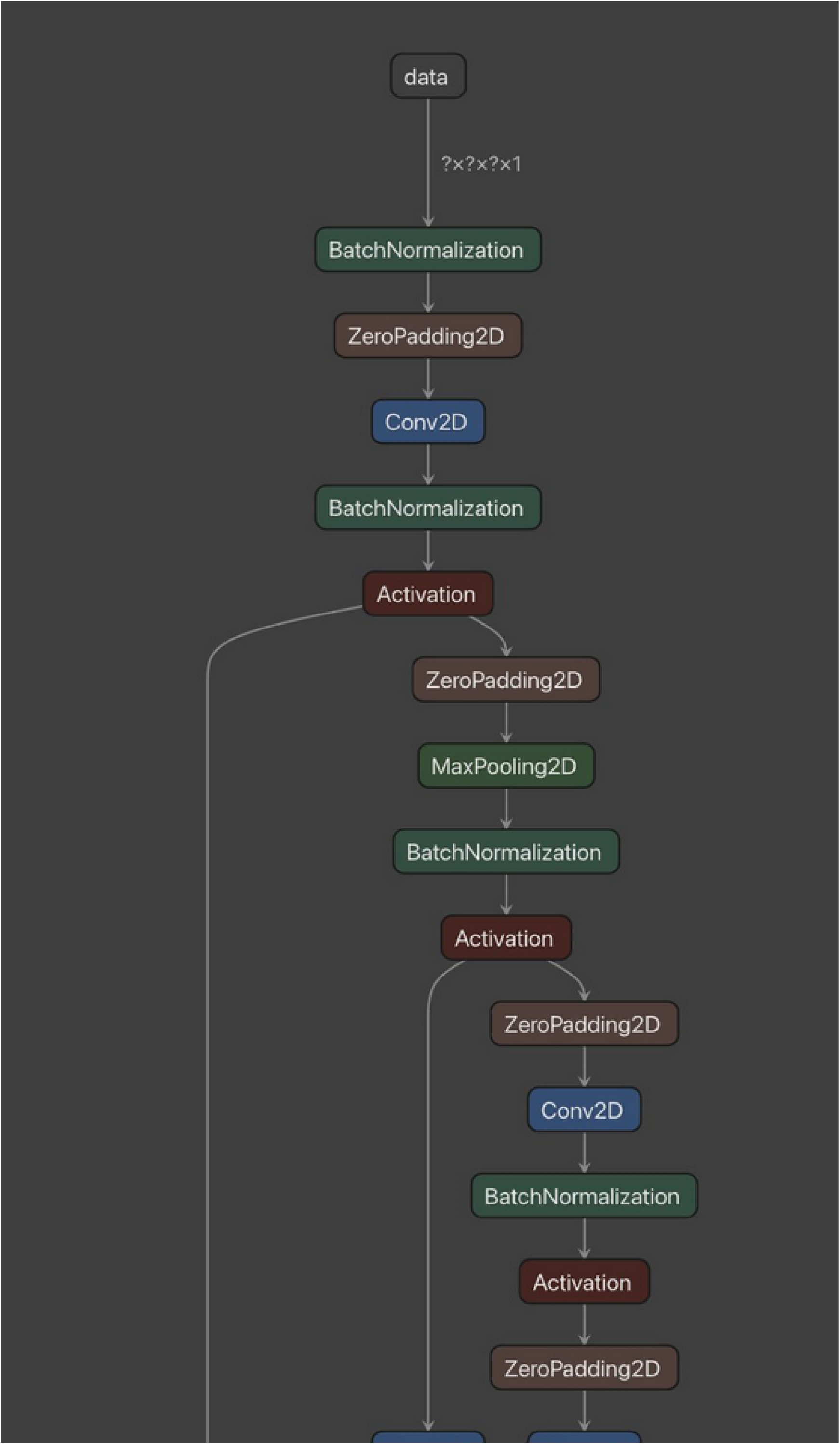
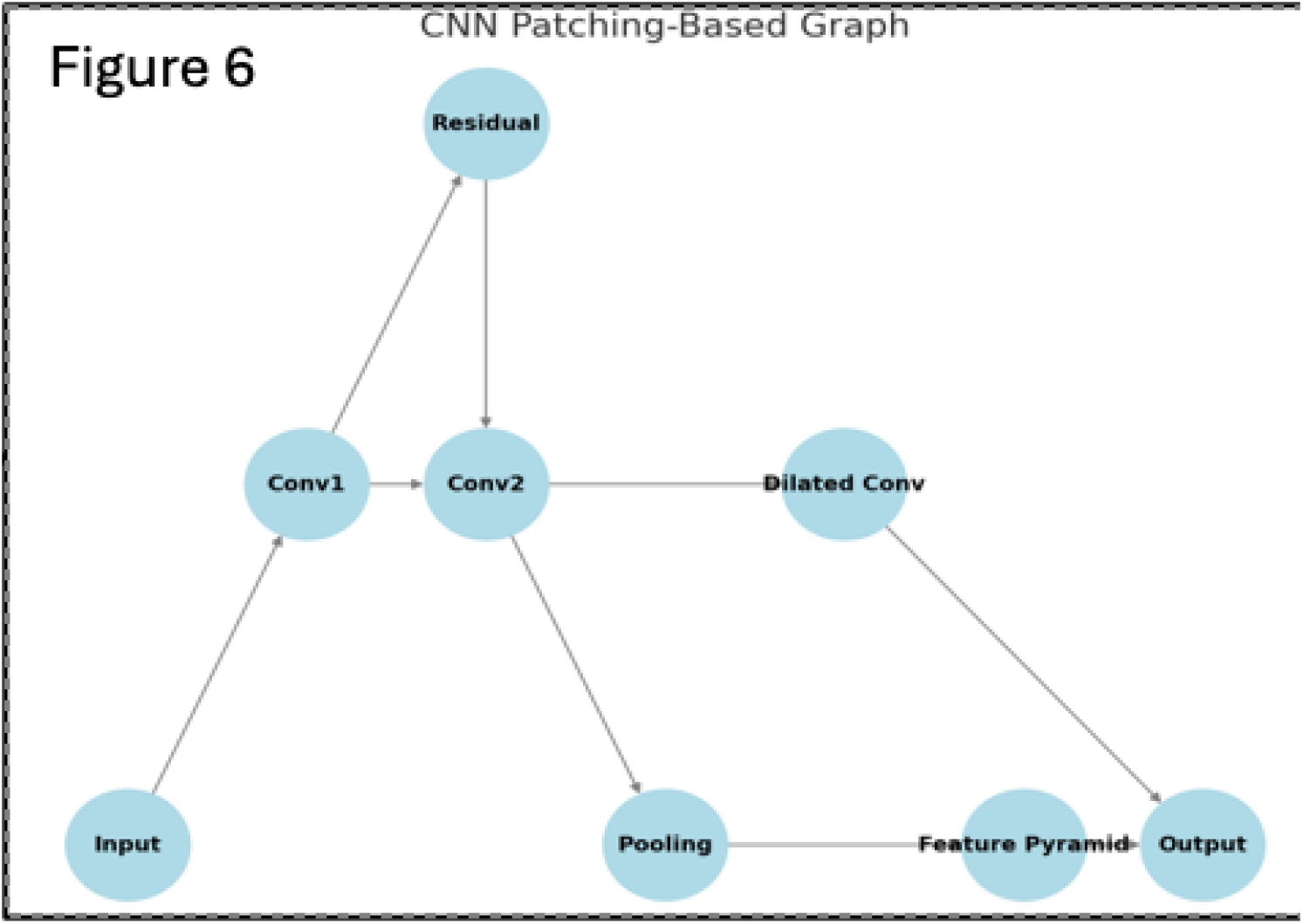

**Figure 2.**
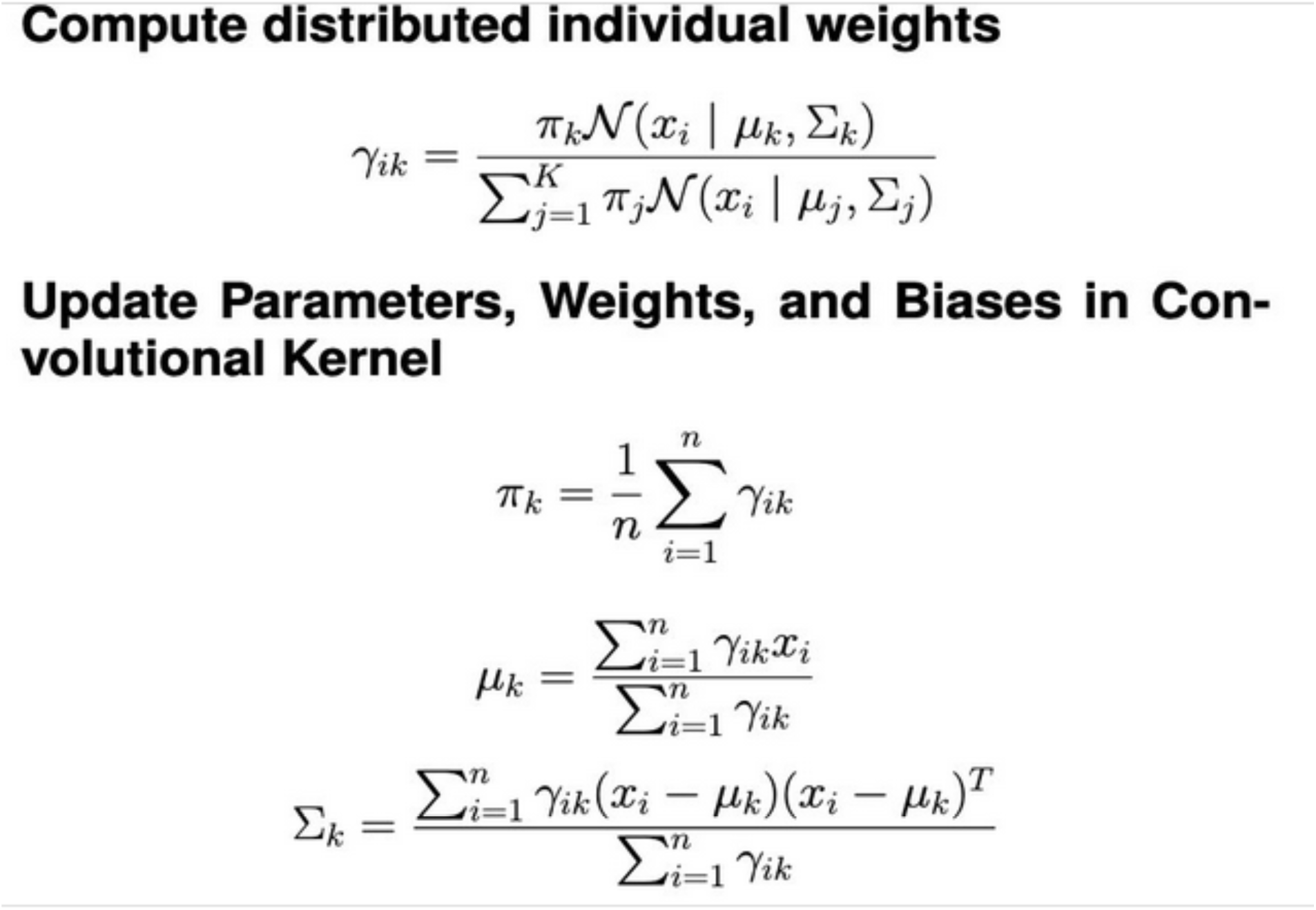
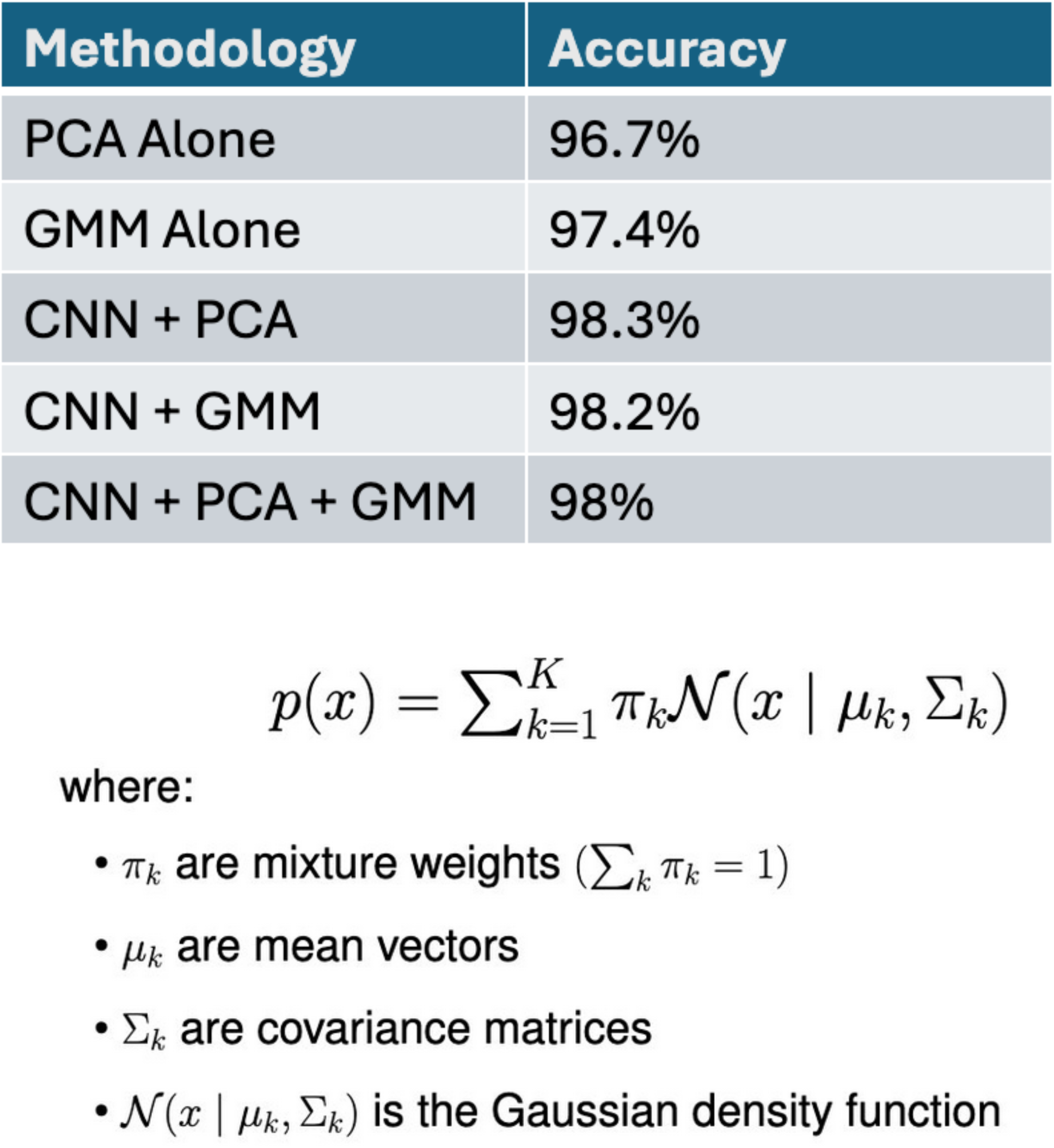
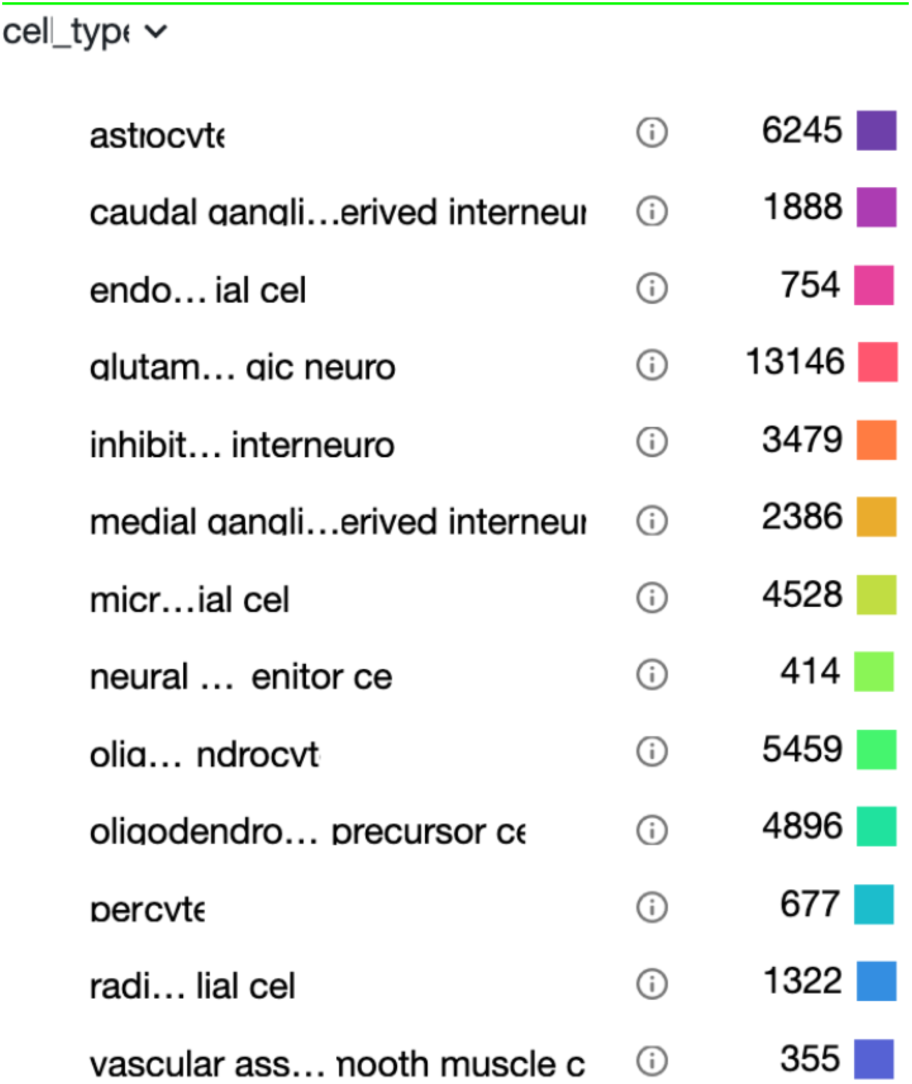

**Figure 3.**
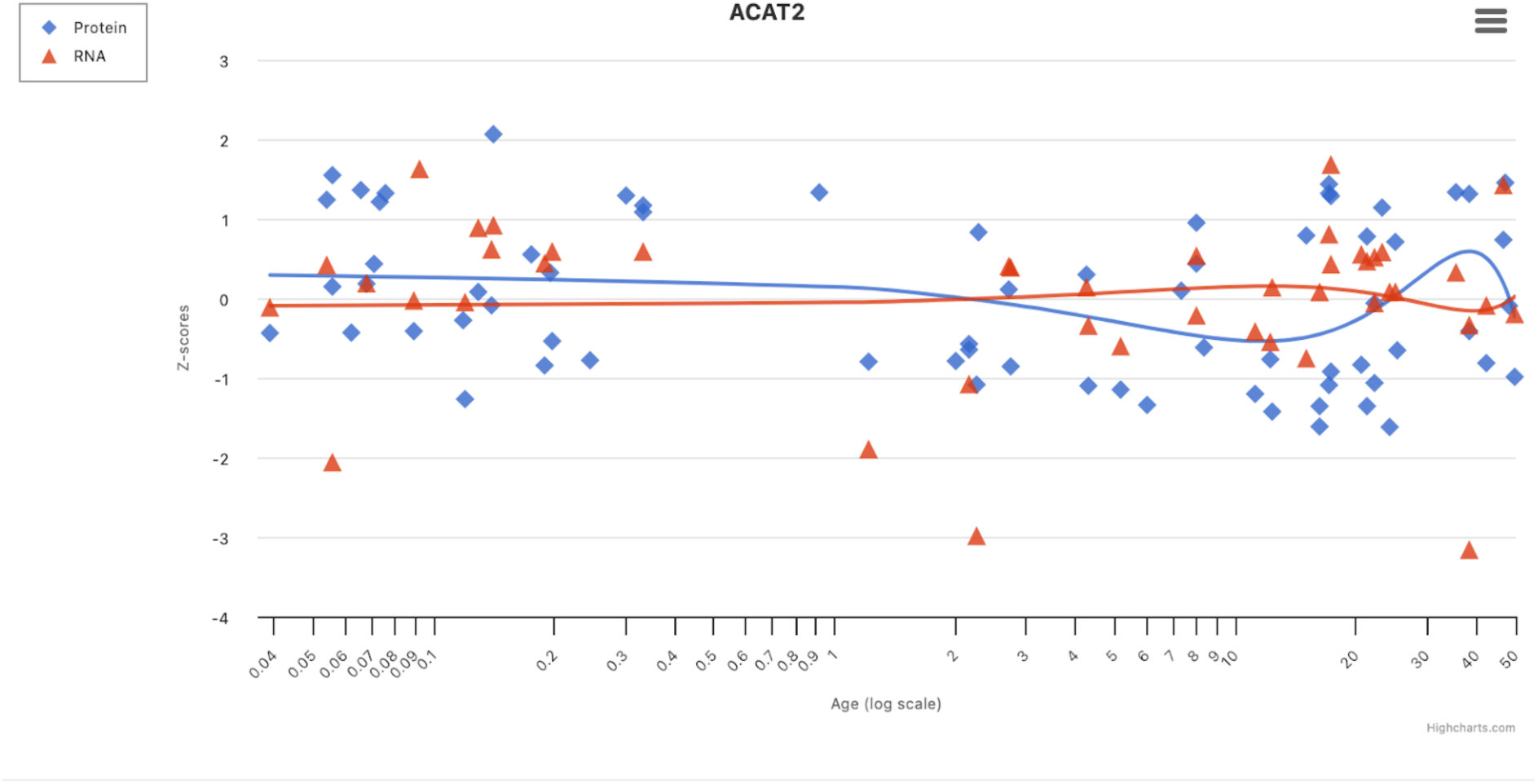

**Figure 4.**
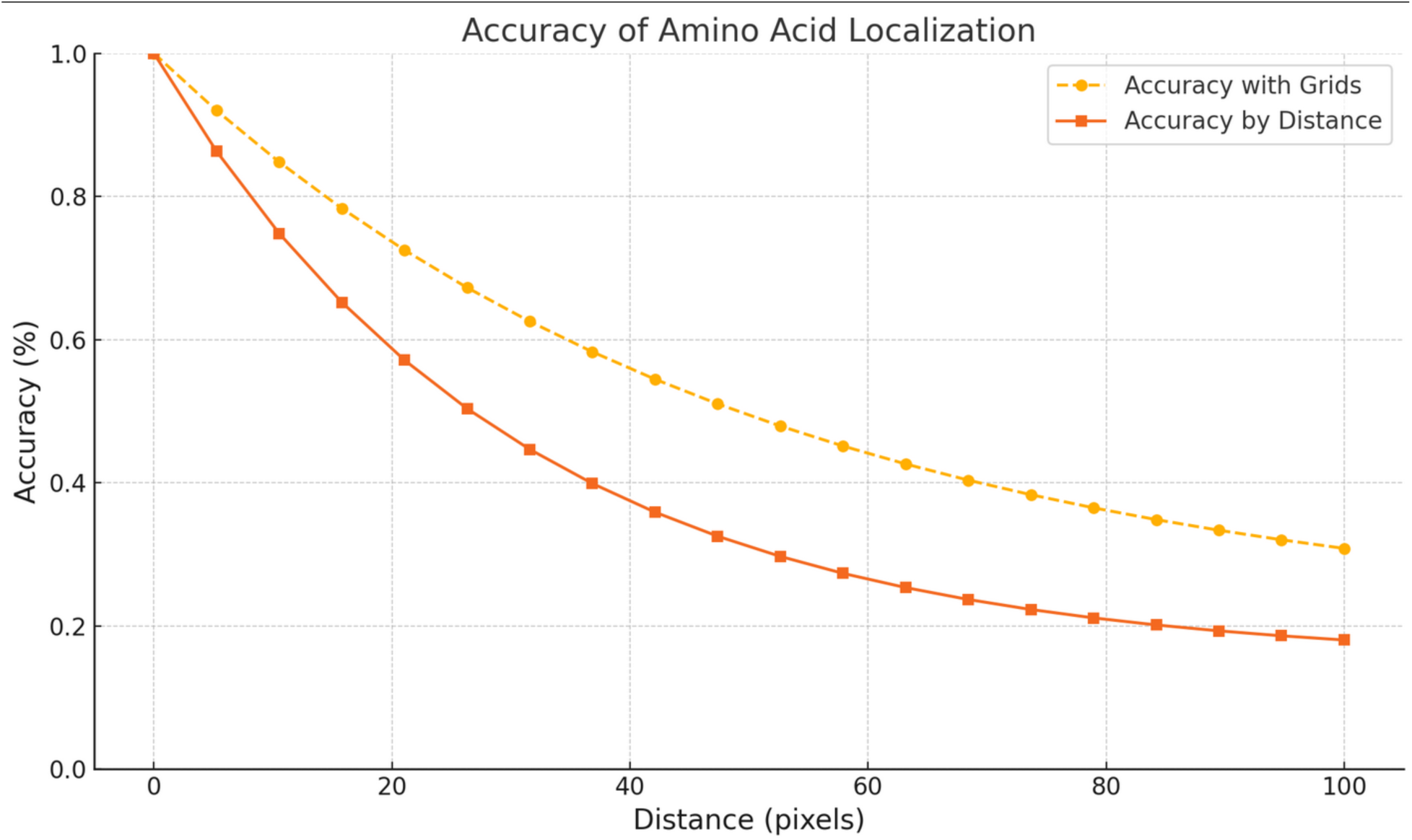

**Figure 5.**
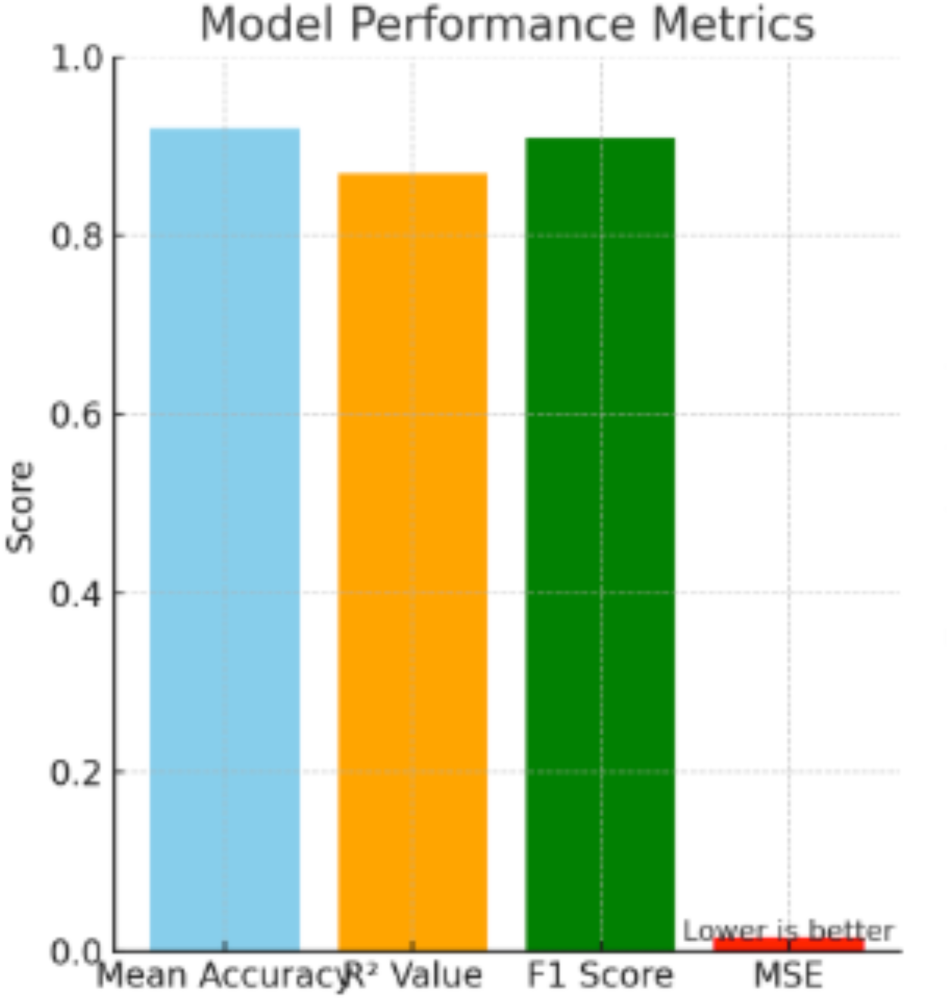

**Figure 6.**
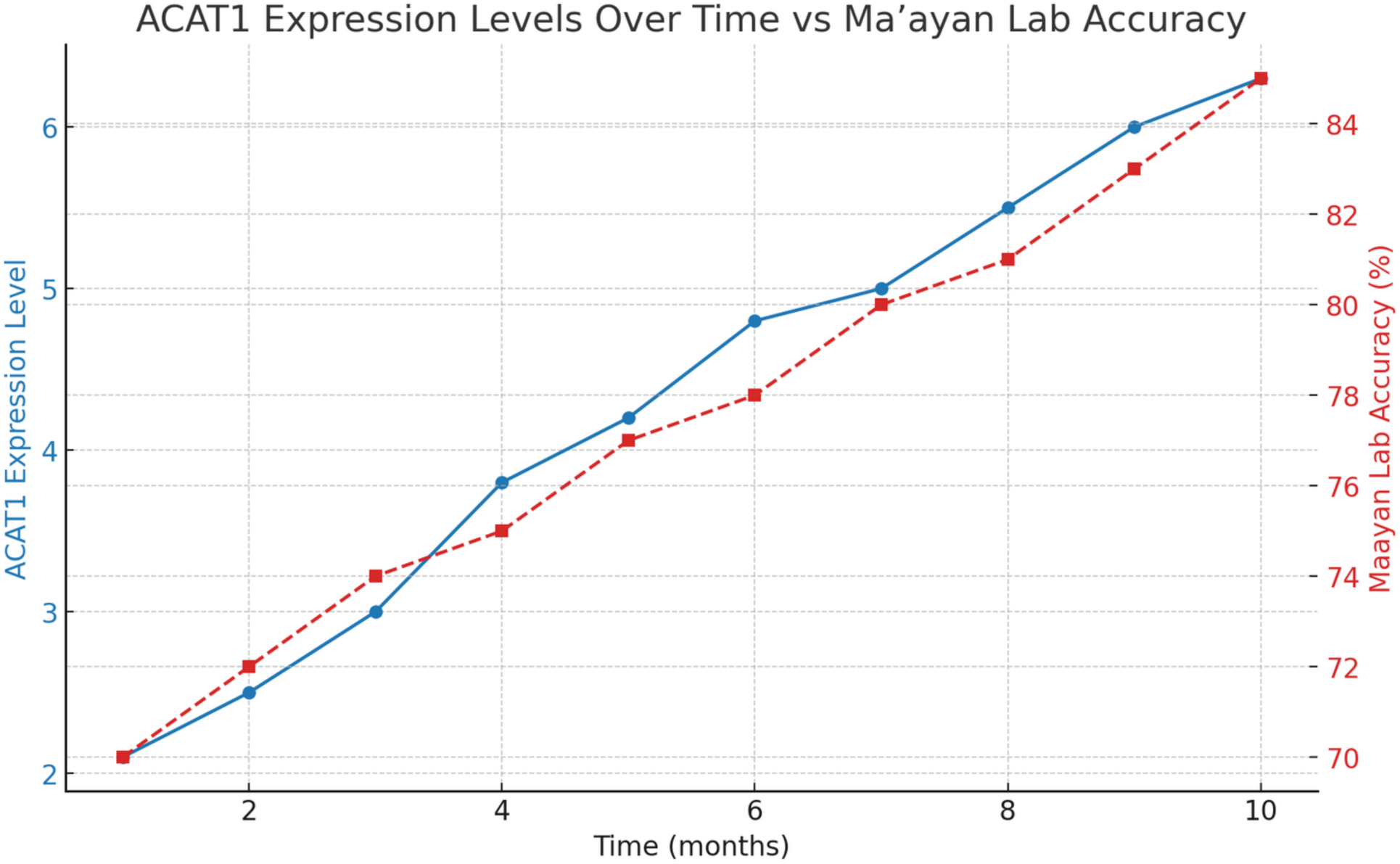

**Figure 7.**
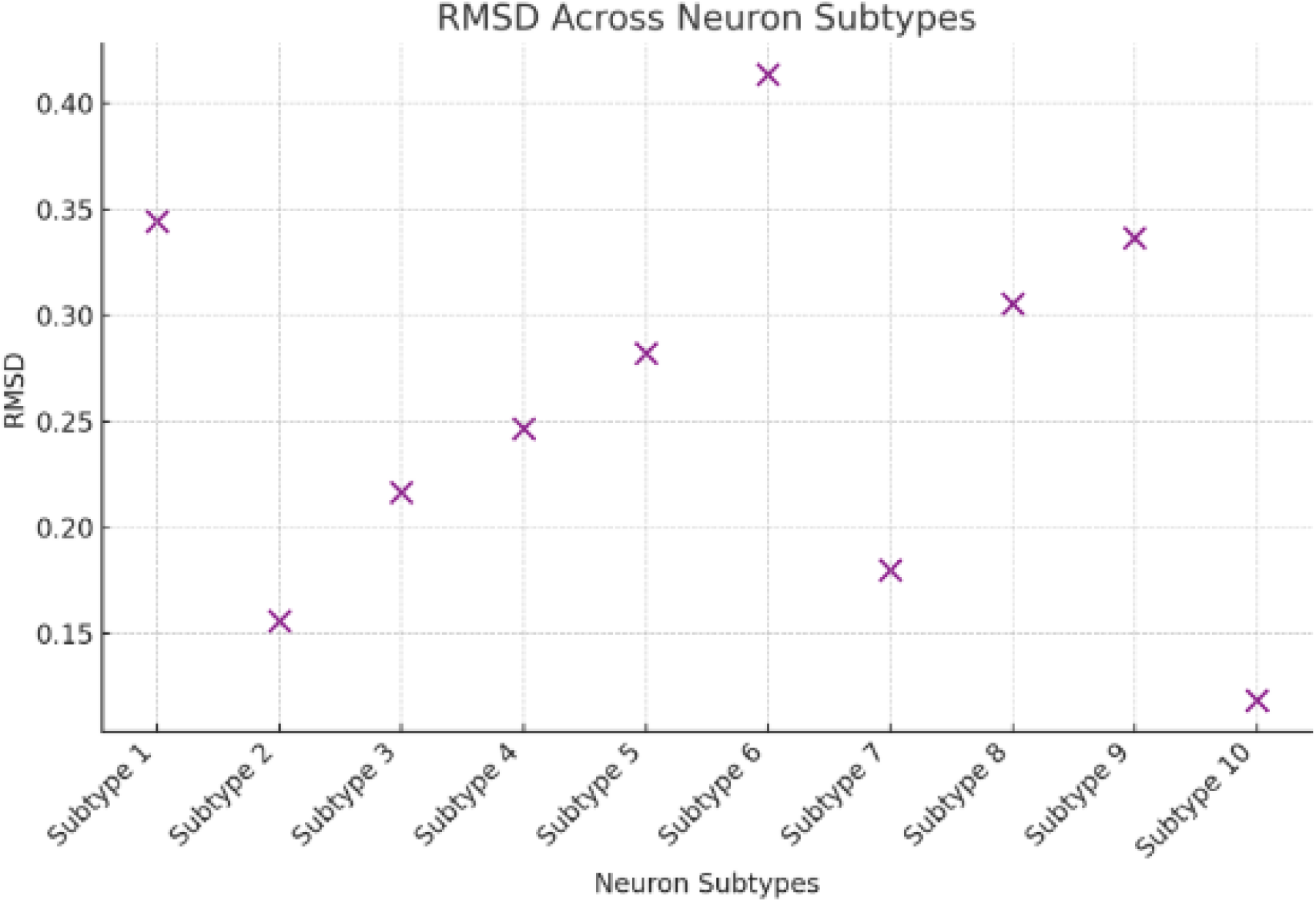

**Figure 8.**
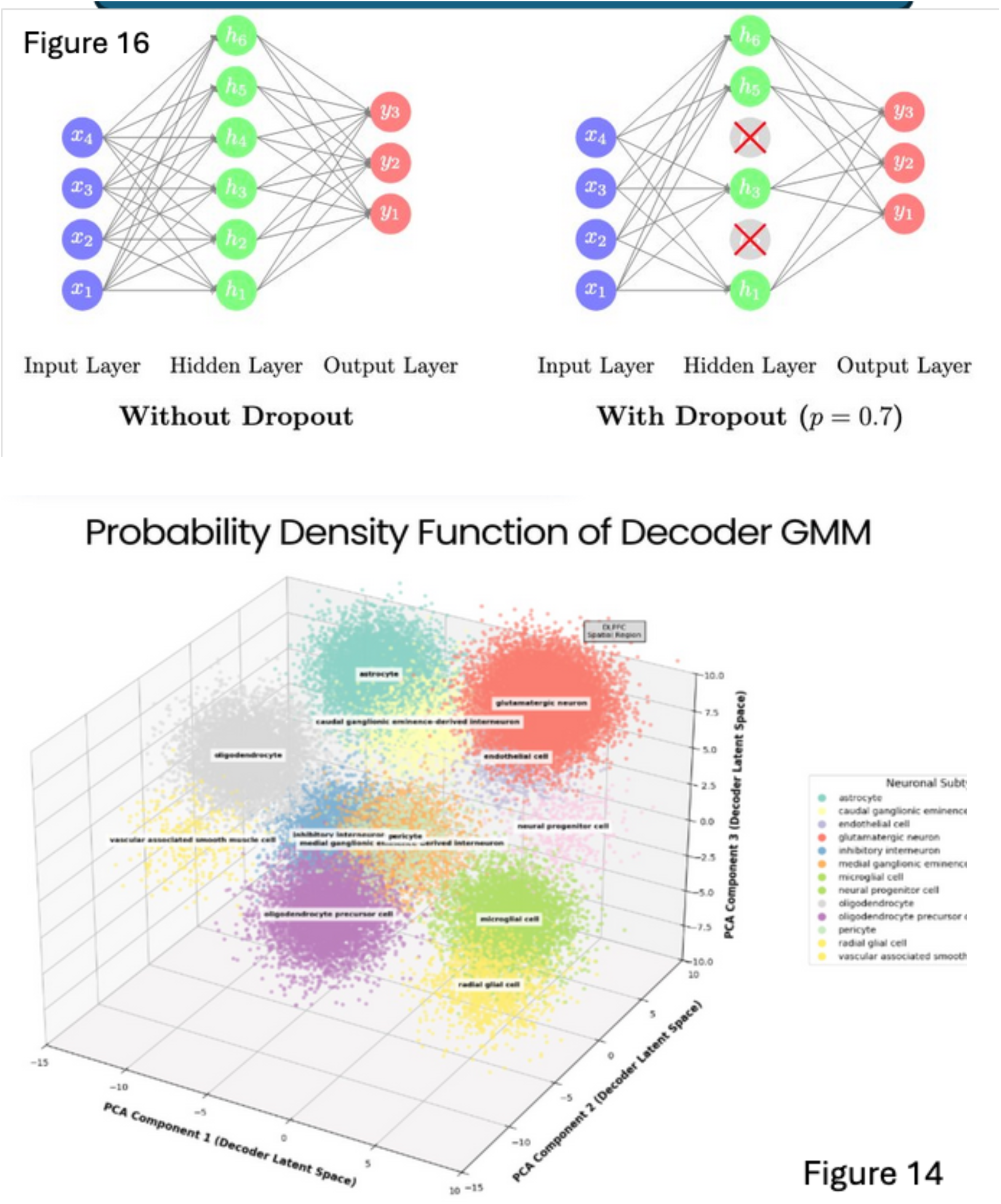
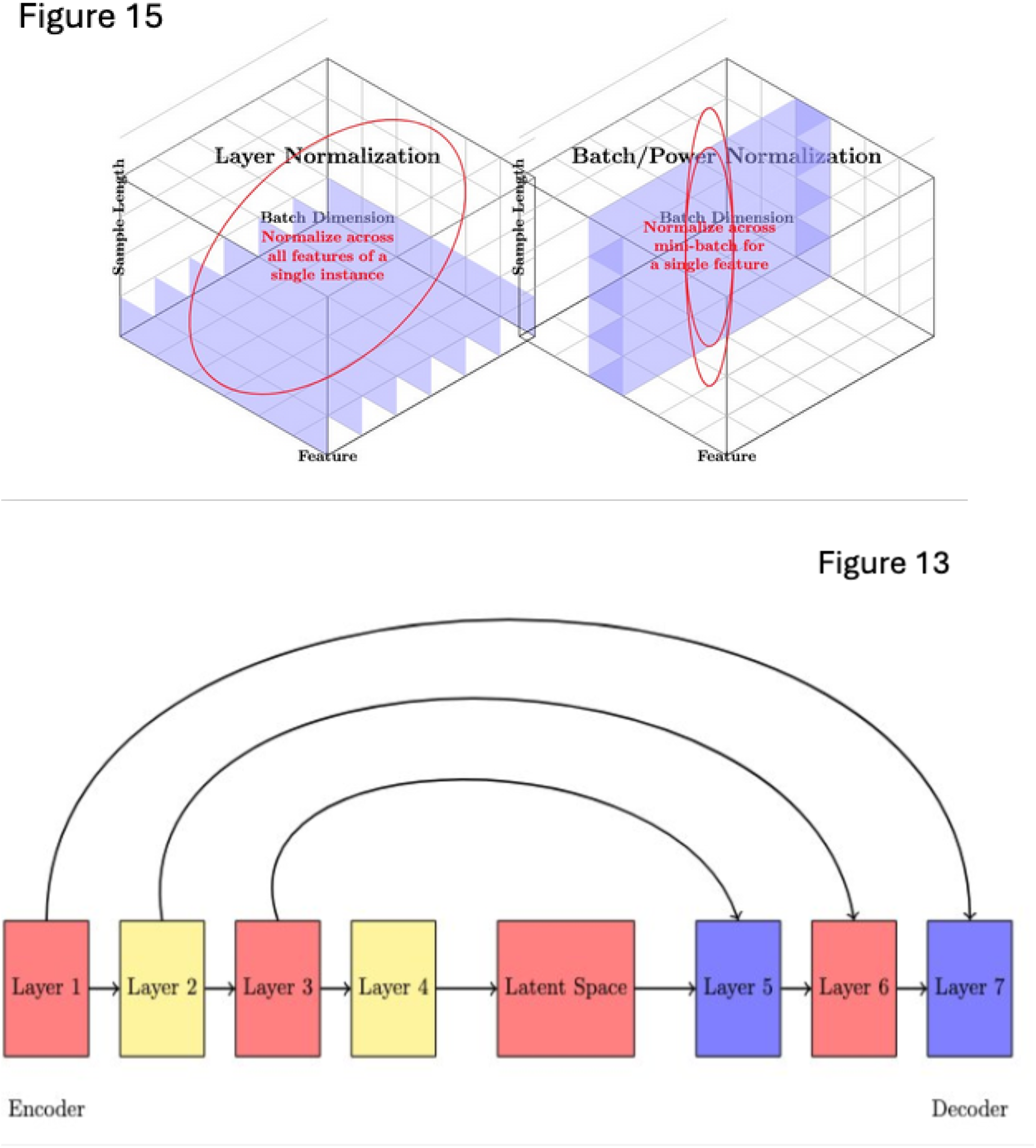
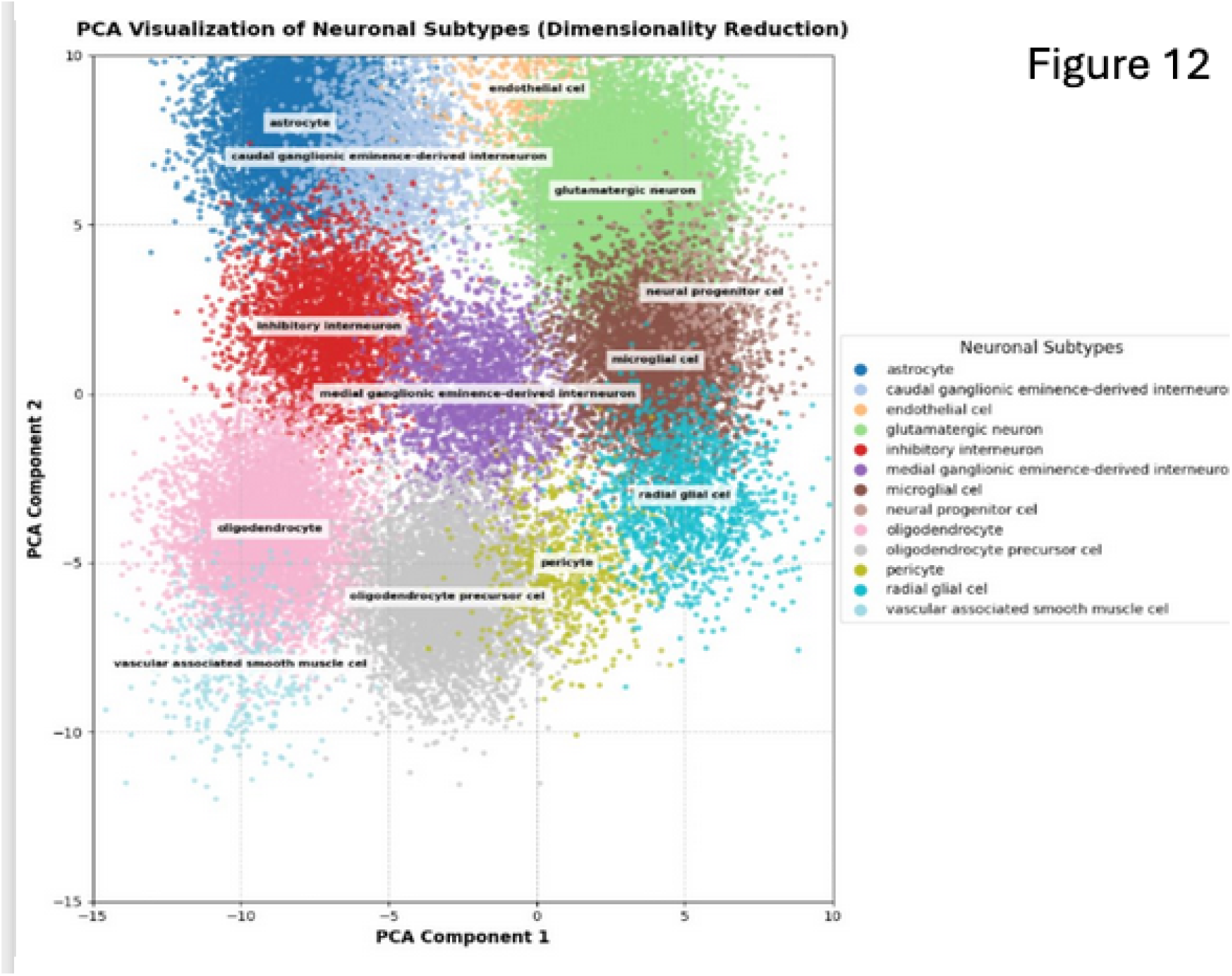
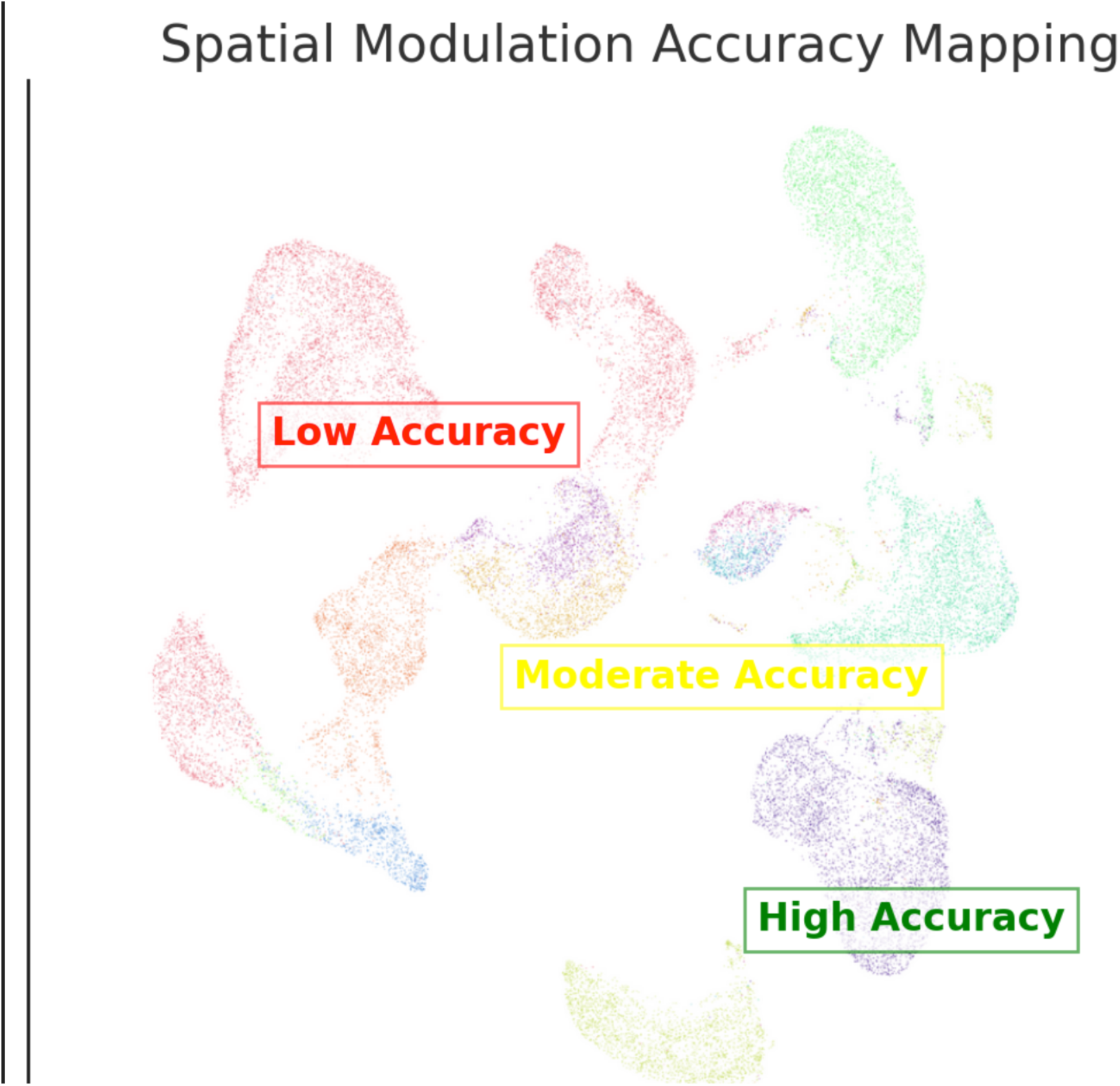

## Supporting information

All Code used in building Sculpt v0.1

## Data Availability

All data produced in the present study are available upon reasonable request to the authors

https://github.com/g4nesh/Sculpt

## Abbreviations

CNN: Convolutional Neural Network
RNN: Recurrent Neural Network
DFLPC: Dorsolateral Prefrontal Cortex

