## Supplementary material for "A Novel Multi-Omics Deep Learning Framework for Spatiotemporal Cerebral Cortex Localization & Expression": All Code used in building Sculpt v0.1: accuracy copy.py

```
import pandas as pd
import numpy as np
from sklearn.model_selection import KFold
from sklearn.preprocessing import StandardScaler
from sklearn.metrics import accuracy_score, precision_score, recall_score, f1_score


def load_database(file_path):
    try:
        return pd.read_csv(file_path)
    except Exception as e:
        raise FileNotFoundError(f"Error loading file: {e}")


def divide_into_segments(sequence, segment_length=10):
    return [sequence[i:i + segment_length] for i in range(0, len(sequence), segment_length)]


def calculate_segment_accuracy(user_sequence, target_sequence, segment_length=10):
    user_segments = divide_into_segments(user_sequence, segment_length)
    target_segments = divide_into_segments(target_sequence, segment_length)

    total_matches = 0
    total_compared = 0

    for user_segment, target_segment in zip(user_segments, target_segments):
        matches = sum(1 for u, t in zip(user_segment, target_segment) if u == t)
        total_matches += matches
        total_compared += len(target_segment)

    accuracy = (total_matches / total_compared) * 100 if total_compared > 0 else 0
    return total_matches, total_compared, accuracy


def z_score_normalization(sequence, mean, std):
    return (sequence - mean) / std


def compute_top_matches(user_sequence, database, top_n=3):
    results = []
    for _, row in database.iterrows():
        total_matches, total_compared, accuracy = calculate_segment_accuracy(
            user_sequence, row['Sequence']
        )
        results.append({
            'Gene': row['Gene'],
            'Sequence': row['Sequence'],
            'Total Matches': total_matches,
            'Total Compared': total_compared,
            'Accuracy (%)': round(accuracy, 2)
        })

    results_df = pd.DataFrame(results).sort_values(by='Accuracy (%)', ascending=False)
    return results_df.head(top_n)


def evaluate_model(database, n_splits=5):
    kf = KFold(n_splits=n_splits, shuffle=True, random_state=42)

    accuracies = []
    precisions = []
    recalls = []
    f1_scores = []

    for train_index, test_index in kf.split(database):
        train_data, test_data = database.iloc[train_index], database.iloc[test_index]

        mean = np.mean(train_data['Sequence'])
        std = np.std(train_data['Sequence'])
        train_data['Normalized Sequence'] = z_score_normalization(train_data['Sequence'], mean, std)
        test_data['Normalized Sequence'] = z_score_normalization(test_data['Sequence'], mean, std)

        y_true = test_data['Gene']
        y_pred = []

        for _, row in test_data.iterrows():
            user_sequence = row['Normalized Sequence']
            top_matches = compute_top_matches(user_sequence, train_data)
            predicted_gene = top_matches.iloc[0]['Gene']
            y_pred.append(predicted_gene)

        accuracies.append(accuracy_score(y_true, y_pred))
        precisions.append(precision_score(y_true, y_pred, average='weighted'))
        recalls.append(recall_score(y_true, y_pred, average='weighted'))
        f1_scores.append(f1_score(y_true, y_pred, average='weighted'))

    print(f"Accuracy: {np.mean(accuracies):.4f}")
    print(f"Precision: {np.mean(precisions):.4f}")
    print(f"Recall: {np.mean(recalls):.4f}")
    print(f"F1 Score: {np.mean(f1_scores):.4f}")


def main():
    file_path = input("Enter the path to your dataset CSV file: ").strip()

    try:
        database = load_database(file_path)
    except FileNotFoundError as e:
        print(e)
        return

    required_columns = {'Gene', 'Sequence'}
    if not required_columns.issubset(database.columns):
        print("Error: The dataset must contain 'Gene' and 'Sequence' columns.")
        return

    user_sequence = input("Enter the amino acid sequence: ").strip().upper()

    if not user_sequence.isalpha():
        print("Error: Sequence must only contain alphabetical characters representing amino acids.")
        return

    try:
        top_matches = compute_top_matches(user_sequence, database)
        print("\nTop Matches with Locality-Based Accuracy:")
        print(top_matches.to_string(index=False))
        evaluate_model(database)
    except Exception as e:
        print(f"Error during analysis: {e}")


if __name__ == "__main__":
    main()
```
