## Supplementary material for "A Novel Multi-Omics Deep Learning Framework for Spatiotemporal Cerebral Cortex Localization & Expression": All Code used in building Sculpt v0.1: BackbonedUNetModel copy.py

```
import Parameter as k2p
from DeepLearning.BackEnd.DeepLearningPipelines.Segmentation.SegmentationModelCreator import SegmentationModelCreator
from DeepLearning.BackEnd.Interface import ModelInterface
from Utils.Common.utils import count_upsampling_layers

#
# "segmentation-models Unet" model factory
# Derived from https://segmentation-models.readthedocs.io/en/latest/api.html#unet
#
"""
Unet is a fully convolution neural network for image semantic segmentation
---
Parameters:
-----------
    backbone_name: name of classification model (without last dense layers) used as feature extractor to build
        segmentation model.
    input_shape: shape of input data/image (H, W, C), in general case you do not need to set H and W shapes, just pass
        (None, None, C) to make your model be able to process images af any size, but H and W of input images should be
        divisible by factor 32.
    classes: a number of classes for output (output shape - (h, w, classes)).
    activation: name of one of keras.activations for last model layer (e.g. sigmoid, softmax, linear).
    weights: optional, path to model weights.
    encoder_weights: one of None (random initialization), imagenet (pre-training on ImageNet).
    encoder_freeze: if True set all layers of encoder (backbone model) as non-trainable.
    encoder_features: a list of layer numbers or names starting from top of the model. Each of these layers will be
        concatenated with corresponding decoder block. If default is used layer names are taken from
        DEFAULT_SKIP_CONNECTIONS.
    decoder_block_type: one of blocks with following layers structure:
    upsampling: UpSampling2D -> Conv2D -> Conv2D
    transpose: Transpose2D -> Conv2D
    decoder_filters: list of numbers of Conv2D layer filters in decoder blocks
    decoder_use_batchnorm: if True, BatchNormalisation layer between Conv2D and Activation layers is used.
"""


class BackbonedUNetModel(SegmentationModelCreator.BaseModel, register_name="BackbonedUNet"):
    def get_parameters_descriptor(bself):
        return k2p.ParameterContainer(
            sub_parameters=(
                k2p.Parameter(
                    name="OutputClassNumber",
                    type=int,
                    validation_policy=k2p.ClampValidationPolicy(2, 1000),
                    default_value=2,
                ),
                k2p.ParameterChoice(name="Backbone", choices=["resnet18", "vgg16", "vgg19"], default_value=0),
            ),
        )

    def __call__(bself, parameters, input_shape=(None, None, 1)):
        if len(input_shape) - 1 == 2:
            import segmentation_models as sm
        else:
            import segmentation_models_3D as sm

        class Ret(ModelInterface):
            def __init__(iself):
                iself._backbone = parameters["Backbone"]
                iself._output_class_nbr = parameters["OutputClassNumber"]
                iself._network_type = "Unet"
                iself._network = None
                if iself._output_class_nbr == 2:
                    iself._activation = "sigmoid"
                    iself._output_class_nbr = 1
                else:
                    iself._activation = "softmax"

                iself._n_dim = len(input_shape) - 1

                # None = random initialisation; imagenet=pretrained weight on imagenet
                iself._encoder_weights = None

                # For now, we do not want to enable this option which load pretrained weights. We can only process
                # grey-level images
                # and LeNet is a rgb public database where the weights have been learned.
                # k2 parameter definition: k2p.Parameter(name="UseLeNet", type=bool, default_value=False),
                # if parameters['UseLeNet']:
                #    nb_channels = 3
                #    encoder_weights = 'imagenet'

                # encoder_freeze: if True set all layers of encoder (backbone model) as non-trainable.
                iself._encoder_freeze = False

            def __call__(iself):
                iself._network = iself._build_network(iself._encoder_weights)
                return iself._network

            def _build_network(iself, encoder_weights):
                return sm.Unet(
                    backbone_name=iself._backbone,
                    classes=iself._output_class_nbr,
                    input_shape=input_shape,
                    encoder_weights=encoder_weights,
                    activation=iself._activation,
                    encoder_freeze=iself._encoder_freeze,
                )

            def get_model_information(iself):
                model = iself._build_network(None)  # None to not load weights: useless here
                multiple_factor = pow(2, count_upsampling_layers(model))
                return iself._network, iself._network_type, iself._backbone, multiple_factor, iself._output_class_nbr

            def get_output_channel_number(iself):
                return iself._output_class_nbr

        return Ret()
```
