## Supplementary material for "A Novel Multi-Omics Deep Learning Framework for Spatiotemporal Cerebral Cortex Localization & Expression": All Code used in building Sculpt v0.1: cnn copy.py

```
import os
import torch
import torch.nn as nn
from torchvision import transforms
from torch.utils.data import DataLoader, Dataset
from PIL import Image
import torch.optim as optim
import numpy as np
import torch.nn.functional as F
import random
import matplotlib.pyplot as plt
import cv2

try:
    import segmentation_models_pytorch as smp
    from torchmetrics.classification import MulticlassJaccardIndex
except ImportError:
    import sys
    sys.path.append(rf'C:\Users\{os.getlogin()}\AppData\Roaming\Python\Python311\site-packages')
    import segmentation_models_pytorch as smp
    from torchmetrics.classification import MulticlassJaccardIndex

# Define the transformation
transform = transforms.Compose([
    transforms.ToTensor(),
    transforms.Normalize(mean=[0.5], std=[0.5])
])

# Function to display a grid of images
def show_images_grid(images, labels, class_names):
    fig, axes = plt.subplots(4, 4, figsize=(8, 8))
    for i, ax in enumerate(axes.flat):
        if i < len(images):
            img = images[i].squeeze().cpu().numpy()
            ax.imshow(img, cmap='gray')
            ax.set_title(f"{class_names[labels[i]]}")
            ax.axis("off")
    plt.show()

# Custom Dataset Class
class NeuronalDataset(Dataset):
    def __init__(self, root_dir, transform=None):
        self.root_dir = root_dir
        self.transform = transform
        self.data = []
        self.class_names = sorted(os.listdir(root_dir))
        for label, class_dir in enumerate(self.class_names):
            class_path = os.path.join(root_dir, class_dir)
            for file in os.listdir(class_path):
                img_path = os.path.join(class_path, file)
                self.data.append((img_path, label))

    def __len__(self):
        return len(self.data)

    def __getitem__(self, idx):
        img_path, label = self.data[idx]
        img = cv2.imread(img_path, cv2.IMREAD_GRAYSCALE)
        img = cv2.resize(img, (128, 128))

        # Convert to tensor and normalize
        if self.transform:
            img = transform(img).float()  # Shape: (1, H, W) -> (1, 128, 128)

        # Add depth dimension for 3D convolution
        img = img.unsqueeze(0)  # Shape: (1, 1, 128, 128)
        img = img.expand(1, 32, 128, 128)  # Shape: (1, D=32, H=128, W=128)

        return img, label

class JaccardLoss(nn.Module):
    def __init__(self, num_classes, smooth=1e-6):
        super(JaccardLoss, self).__init__()
        self.num_classes = num_classes
        self.smooth = smooth

    def forward(self, inputs, targets):
        inputs = F.softmax(inputs, dim=1)
        targets_one_hot = F.one_hot(targets, num_classes=self.num_classes).float()

        intersection = (inputs * targets_one_hot).sum(dim=1)
        union = inputs.sum(dim=1) + targets_one_hot.sum(dim=1) - intersection

        iou = (intersection + self.smooth) / (union + self.smooth)
        return 1 - iou.mean()

class UNet(nn.Module):
    def __init__(self, input_channels=1, output_classes=5):
        super(UNet, self).__init__()

        self.encoder = nn.Sequential(
            nn.Conv3d(input_channels, 64, kernel_size=3, padding=1),
            nn.BatchNorm3d(64),
            nn.ReLU(inplace=True),
            nn.MaxPool3d(kernel_size=2, stride=2)
        )

        self.middle = nn.Sequential(
            nn.Conv3d(64, 128, kernel_size=3, padding=1),
            nn.BatchNorm3d(128),
            nn.ReLU(inplace=True),
            nn.Conv3d(128, 128, kernel_size=3, padding=1),
            nn.BatchNorm3d(128),
            nn.ReLU(inplace=True)
        )

        self.decoder = nn.Sequential(
            nn.Conv3d(128, 64, kernel_size=3, padding=1),
            nn.BatchNorm3d(64),
            nn.ReLU(inplace=True),
            nn.ConvTranspose3d(64, 64, kernel_size=2, stride=2)  # Upsamples D, H, W
        )

        self.pool = nn.AdaptiveAvgPool3d(1)
        self.final = nn.Conv3d(64, output_classes, kernel_size=1)

    def forward(self, x):
        x1 = self.encoder(x)  # Shape: (batch_size, 64, 16, 64, 64)
        x2 = self.middle(x1)  # Shape: (batch_size, 128, 16, 64, 64)
        x3 = self.decoder(x2)  # Shape: (batch_size, 64, 32, 128, 128)

        # Global pooling and final classification
        x4 = self.pool(x3)  # Shape: (batch_size, 64, 1, 1, 1)
        out = self.final(x4)  # Shape: (batch_size, output_classes, 1, 1, 1)
        out = out.squeeze(-1).squeeze(-1).squeeze(-1)  # Shape: (batch_size, output_classes)

        return out

# Training Function
def train_model(root_dir, num_epochs=10, batch_size=4, learning_rate=0.001):
    # Setup device
    device = torch.device("cuda" if torch.cuda.is_available() else "cpu")
    print(f"Using device: {device}")

    # Create dataset and dataloader
    dataset = NeuronalDataset(root_dir=root_dir, transform=transform)
    dataloader = DataLoader(dataset, batch_size=batch_size, shuffle=True)

    # Initialize model
    model = UNet(input_channels=1, output_classes=5).to(device)

    # Loss and optimizer
    criterion_ce = nn.CrossEntropyLoss()
    criterion_jaccard = JaccardLoss(num_classes=5)
    optimizer = optim.Adam(model.parameters(), lr=learning_rate)

    # Metrics
    jaccard_metric = MulticlassJaccardIndex(num_classes=5).to(device)

    # Training loop
    for epoch in range(num_epochs):
        model.train()
        total_loss = 0
        total_iou = 0
        correct = 0
        total = 0

        for images, labels in dataloader:
            images = images.to(device)  # Shape: (batch_size, 1, 32, 128, 128)
            labels = labels.to(device)  # Shape: (batch_size)

            # Zero gradients
            optimizer.zero_grad()

            # Forward pass
            outputs = model(images)  # Shape: (batch_size, 5)

            # Compute losses
            loss_ce = criterion_ce(outputs, labels)
            loss_jaccard = criterion_jaccard(outputs, labels)
            loss = loss_ce + loss_jaccard  # Combine losses

            # Backward pass and optimize
            loss.backward()
            optimizer.step()

            # Statistics
            total_loss += loss.item()
            _, predicted = torch.max(outputs, 1)  # Shape: (batch_size)
            total += labels.size(0)
            correct += (predicted == labels).sum().item()

            # Calculate Jaccard index (IoU)
            jaccard_score = jaccard_metric(outputs, labels)
            total_iou += jaccard_score.item()

        # Epoch statistics
        accuracy = 100 * correct / total
        avg_loss = total_loss / len(dataloader)
        avg_iou = total_iou / len(dataloader)

        print(f"Epoch [{epoch + 1}/{num_epochs}]")
        print(f"Loss: {avg_loss:.4f} (CE + Jaccard)")
        print(f"Accuracy: {accuracy:.2f}%")
        print(f"IoU Score: {avg_iou:.4f}")
        print("-" * 50)

    return model

# Prediction Function
def predict(model, image_path, device):
    model.eval()

    # Load and preprocess image
    img = cv2.imread(image_path, cv2.IMREAD_GRAYSCALE)
    img = cv2.resize(img, (128, 128))

    # Transform
    if transform:
        img = transform(img).float()  # Shape: (1, 128, 128)

    # Add depth dimension
    img = img.unsqueeze(0)  # Shape: (1, 1, 128, 128)
    img = img.expand(1, 32, 128, 128)  # Shape: (1, 32, 128, 128)
    img = img.unsqueeze(0)  # Shape: (1, 1, 32, 128, 128)

    img = img.to(device)

    with torch.no_grad():
        output = model(img)  # Shape: (1, 5)
        _, predicted = torch.max(output, 1)  # Shape: (1)

    return predicted.item()

# Main Execution
if __name__ == "__main__":
    # Set random seeds for reproducibility
    torch.manual_seed(42)
    np.random.seed(42)
    random.seed(42)

    # Training parameters
    ROOT_DIR = "/Users/ganeshtalluri/PycharmProjects/Sculpt/Patches"  # Adjust this path
    NUM_EPOCHS = 10
    BATCH_SIZE = 4
    LEARNING_RATE = 0.001

    # Train model
    model = train_model(
        root_dir=ROOT_DIR,
        num_epochs=NUM_EPOCHS,
        batch_size=BATCH_SIZE,
        learning_rate=LEARNING_RATE
    )

    # Save model
    torch.save(model.state_dict(), "neuronal_unet_model.pth")
    print("Model saved to 'neuronal_unet_model.pth'")

    # Load a sample image and make prediction
    dataset = NeuronalDataset(root_dir=ROOT_DIR, transform=transform)
    sample_img_path, _ = dataset.data[0]
    device = torch.device("cuda" if torch.cuda.is_available() else "cpu")
    prediction = predict(model, sample_img_path, device)
    print(f"Prediction for sample image: {dataset.class_names[prediction]}")

    # Show sample images
    dataloader = DataLoader(dataset, batch_size=16, shuffle=True)
    sample_images, sample_labels = next(iter(dataloader))
    show_images_grid(sample_images[:, :, 16, :, :], sample_labels, dataset.class_names)  # Show middle slice
```
