## Supplementary material for "A Novel Multi-Omics Deep Learning Framework for Spatiotemporal Cerebral Cortex Localization & Expression": All Code used in building Sculpt v0.1: conjoined copy.py

```
import torch
import torch.nn as nn
import torch.optim as optim
import torchvision.transforms as transforms
from torch.utils.data import DataLoader, Dataset
import cv2
import os
import numpy as np

class NeuronalDataset(Dataset):
    def __init__(self, root_dir, transform=None, seq_len=4, num_classes=5):
        self.root_dir = root_dir
        self.transform = transform
        self.seq_len = seq_len
        self.num_classes = num_classes
        self.data = []

        for label, class_dir in enumerate(sorted(os.listdir(root_dir))):
            class_path = os.path.join(root_dir, class_dir)
            for file in os.listdir(class_path):
                img_path = os.path.join(class_path, file)
                self.data.append((img_path, label))

    def __len__(self):
        return len(self.data)

    def __getitem__(self, idx):
        img_path, label = self.data[idx]
        img = cv2.imread(img_path, cv2.IMREAD_GRAYSCALE)
        img = cv2.resize(img, (128, 128))
        img = np.expand_dims(img, axis=0)
        img = img.transpose((1, 2, 0))

        if self.transform:
            img = self.transform(img)

        labels = torch.zeros((self.seq_len, self.num_classes))
        labels[:, label] = 1

        return img, labels

transform = transforms.Compose([
    transforms.ToTensor(),
    transforms.Normalize(mean=[0.5], std=[0.5])
])

dataset = NeuronalDataset(root_dir="/Users/ganeshtalluri/PycharmProjects/Sculpt/Patches", transform=transform)
dataloader = DataLoader(dataset, batch_size=16, shuffle=True)
sample_img, _ = dataset[0]
print("Image shape:", sample_img.shape)

class HybridCNNRNN(nn.Module):
    def __init__(self, num_classes=5, hidden_size=128, num_layers=2):
        super(HybridCNNRNN, self).__init__()

        self.cnn = nn.Sequential(
            nn.Conv2d(1, 32, kernel_size=3, stride=1, padding=1),
            nn.ReLU(),
            nn.MaxPool2d(kernel_size=2, stride=2),

            nn.Conv2d(32, 64, kernel_size=3, stride=1, padding=1),
            nn.ReLU(),
            nn.MaxPool2d(kernel_size=2, stride=2),

            nn.Conv2d(64, 128, kernel_size=3, stride=1, padding=1),
            nn.ReLU(),
            nn.MaxPool2d(kernel_size=2, stride=2)
        )

        self.fc = nn.Linear(128 * 16 * 16, hidden_size)

        self.rnn = nn.LSTM(input_size=hidden_size, hidden_size=hidden_size, num_layers=num_layers, batch_first=True)

        self.output_layer = nn.Linear(hidden_size, num_classes)

    def forward(self, x):
        batch_size, seq_len, channels, height, width = x.size()

        cnn_features = []
        for t in range(seq_len):
            cnn_out = self.cnn(x[:, t, :, :, :])
            cnn_out = cnn_out.view(batch_size, -1)
            cnn_out = self.fc(cnn_out)
            cnn_features.append(cnn_out)

        cnn_features = torch.stack(cnn_features, dim=1)

        rnn_out, _ = self.rnn(cnn_features)

        outputs = self.output_layer(rnn_out)

        return outputs

model = HybridCNNRNN(num_classes=5, hidden_size=128, num_layers=2)
device = torch.device("cuda" if torch.cuda.is_available() else "cpu")
model.to(device)

criterion = nn.MSELoss()
optimizer = optim.Adam(model.parameters(), lr=0.001)

num_epochs = 10
for epoch in range(num_epochs):
    total_loss = 0
    for images, labels in dataloader:
        images, labels = images.to(device), labels.to(device)

        images = images.unsqueeze(1).repeat(1, 4, 1, 1, 1)

        optimizer.zero_grad()
        outputs = model(images)
        loss = criterion(outputs, labels)
        loss.backward()
        optimizer.step()

        total_loss += loss.item()

    print(f"Epoch [{epoch + 1}/{num_epochs}], Loss: {total_loss / len(dataloader):.4f}")

torch.save(model.state_dict(), "hybrid_cnn_rnn.pth")

def predict(model, image_path):
    model.eval()
    image = cv2.imread(image_path, cv2.IMREAD_GRAYSCALE)
    image = cv2.resize(image, (128, 128))
    image = np.expand_dims(image, axis=0)
    image = image.transpose((1, 2, 0))
    image = transform(image).unsqueeze(0).to(device)

    image = image.unsqueeze(1).repeat(1, 4, 1, 1, 1)

    with torch.no_grad():
        output = model(image)
        predictions = output.squeeze().cpu().numpy()

    return predictions
```
