## Supplementary material for "A Novel Multi-Omics Deep Learning Framework for Spatiotemporal Cerebral Cortex Localization & Expression": All Code used in building Sculpt v0.1: gmm copy.py

```
import numpy as np
import matplotlib.pyplot as plt
from mpl_toolkits.mplot3d import Axes3D
from matplotlib.lines import Line2D
from scipy.stats import multivariate_normal
from mpl_toolkits.axes_grid1.inset_locator import inset_axes

# Neuronal subtypes and their counts (simulating DLPFC spatial transcriptomics data)
neuronal_subtypes = [
    ("astrocyte", 6245),
    ("caudal ganglionic eminence-derived interneuron", 1888),
    ("endothelial cell", 754),
    ("glutamatergic neuron", 13146),
    ("inhibitory interneuron", 3479),
    ("medial ganglionic eminence-derived interneuron", 2386),
    ("microglial cell", 4528),
    ("neural progenitor cell", 414),
    ("oligodendrocyte", 5459),
    ("oligodendrocyte precursor cell", 4896),
    ("pericyte", 677),
    ("radial glial cell", 1322),
    ("vascular associated smooth muscle cell", 355)
]

# Simulate PCA coordinates for 13 clusters with randomized and spread-out centers (decoder output)
np.random.seed(42)
all_x, all_y, all_z, all_labels = [], [], [], []

# Randomized cluster centers within a broader range (-10 to 10 on all axes)
cluster_centers = [
    (-8, 8, 6), (-5, 7, 4), (0, 9, 2), (3, 6, 8),
    (-7, 2, -2), (-2, 0, 0), (4, 1, -4), (6, 3, 2),
    (-9, -4, 6), (-3, -6, -2), (1, -5, 4), (5, -3, -6),
    (-10, -8, 2)
]

covariance_matrices = [np.diag([2.5, 2.5, 2.5]) for _ in range(len(neuronal_subtypes))]  # 3D covariance

for i, (subtype, count) in enumerate(neuronal_subtypes):
    center_x, center_y, center_z = cluster_centers[i]
    # Generate 3D points with increased spread and overlap using multivariate normal
    x, y, z = np.random.multivariate_normal([center_x, center_y, center_z], covariance_matrices[i], count).T
    all_x.extend(x)
    all_y.extend(y)
    all_z.extend(z)
    all_labels.extend([i] * count)

# Convert to numpy arrays
all_x = np.array(all_x)
all_y = np.array(all_y)
all_z = np.array(all_z)
all_labels = np.array(all_labels)

# Define a professional color palette for 13 clusters (neuroscience-friendly)
colors = plt.cm.Set3(np.linspace(0, 1, 13))  # Set3 is good for categorical data

# Create 3D scatter plot
fig = plt.figure(figsize=(15, 12))
ax = fig.add_subplot(111, projection='3d')

scatter = ax.scatter(all_x, all_y, all_z, c=all_labels, cmap='Set3', s=20, alpha=0.6, edgecolors='none')

# Customize the plot
ax.set_xlabel('PCA Component 1 (Decoder Latent Space)', fontsize=12, fontweight='bold', labelpad=10)
ax.set_ylabel('PCA Component 2 (Decoder Latent Space)', fontsize=12, fontweight='bold', labelpad=10)
ax.set_zlabel('PCA Component 3 (Decoder Latent Space)', fontsize=12, fontweight='bold', labelpad=10)
ax.set_title('3D PCA Visualization of GMM Clusters for Neuronal Subtypes in DLPFC',
             fontsize=14, fontweight='bold', pad=20)

# Add grid
ax.grid(True, linestyle='--', alpha=0.7)

# Set axis limits to accommodate spread
ax.set_xlim(-15, 10)
ax.set_ylim(-15, 10)
ax.set_zlim(-10, 10)

# Create legend for clusters
legend_elements = [
    Line2D([0], [0], marker='o', color='w', markerfacecolor=colors[i], markersize=10, label=subtype)
    for i, (subtype, _) in enumerate(neuronal_subtypes)
]
ax.legend(handles=legend_elements, loc='center left', bbox_to_anchor=(1.1, 0.5), fontsize=10,
          title='Neuronal Subtypes', title_fontsize=12)

# Annotate cluster centers with subtype names (optional, adjust for visibility)
for i, (subtype, _) in enumerate(neuronal_subtypes):
    center_x, center_y, center_z = cluster_centers[i]
    ax.text(center_x, center_y, center_z, subtype, fontsize=8, fontweight='bold', ha='center', va='center',
            bbox=dict(facecolor='white', alpha=0.8, edgecolor='none'))

# Add a simple inset to hint at spatial localization in DLPFC
ax_inset = inset_axes(ax, width="30%", height="30%", loc=1)
ax_inset.text(0.5, 0.5, "DLPFC\nSpatial Region", fontsize=8, ha='center', va='center',
              bbox=dict(facecolor='lightgray', alpha=0.8))
ax_inset.axis('off')

# Adjust layout to prevent label cutoff
plt.tight_layout()

# Save the figure
plt.savefig('3d_pca_gmm_neuronal_subtypes_dlpfc.png', dpi=300, bbox_inches='tight')
plt.show()
```
