## Supplementary material for "A Novel Multi-Omics Deep Learning Framework for Spatiotemporal Cerebral Cortex Localization & Expression": All Code used in building Sculpt v0.1: maayanlab copy.txt

README, file descriptions.
Matrix_proteomics.txt = matrix of protein abundance (rows = samples; columns=protein IDs)
Meta_proteomics.txt = metadata file for 69 proteome samples
Matrix_transcriptomics.txt = matrix of gene expression (rows = samples; columns=gene IDs)
Meta_transcriptomics.txt = metadata file for 44 transcriptome samples
ModuleLists.gmt = .gmt file containing protein-module and gene-module relationships for both the proteome and transcriptome, respectively.
