## Supplementary material for "A Novel Multi-Omics Deep Learning Framework for Spatiotemporal Cerebral Cortex Localization & Expression": All Code used in building Sculpt v0.1: patching copy.py

```
import cv2
import numpy as np
import os
from skimage.util import view_as_windows
from sklearn.cluster import KMeans

image = cv2.imread("CELLxGENE_umap_emb.png", cv2.IMREAD_GRAYSCALE)
image = image / 255.0

patch_size = (128, 128)
stride = 128

patches = view_as_windows(image, patch_size, step=stride)
patches = patches.reshape(-1, patch_size[0], patch_size[1])

num_patches = patches.shape[0]
patches_flat = patches.reshape(num_patches, -1)

cell_types = [
    "Neural progenitor cell", "Radial glial cell", "Astrocyte",
    "Glutamatergic neuron", "Inhibitory interneuron",
    "Endothelial cell", "Pericyte",
    "Oligodendrocyte", "Oligodendrocyte precursor cell",
    "Microglial cell", "Vascular associated smooth muscle cell",
    "Glutamatergic neuron", "Inhibitory interneuron",
    "Neural progenitor cell", "Glutamatergic neuron", "Inhibitory interneuron", "Astrocyte",
    "Oligodendrocyte", "Oligodendrocyte precursor cell", "Radial glial cell", "Microglial cell"
]

n_clusters = len(cell_types)
kmeans = KMeans(n_clusters=n_clusters, random_state=42, n_init=10)
kmeans.fit(patches_flat)
labels = kmeans.labels_

for cell_type in cell_types:
    os.makedirs(cell_type, exist_ok=True)

for i, (patch, label) in enumerate(zip(patches, labels)):
    cell_type = cell_types[label]
    cv2.imwrite(f"{cell_type}/patch_{i}.png", (patch * 255).astype(np.uint8))

print(f"Total patches: {num_patches}")
print(f"Patches grouped into {n_clusters} cell types.")
```
