## Supplementary material for "A Novel Multi-Omics Deep Learning Framework for Spatiotemporal Cerebral Cortex Localization & Expression": All Code used in building Sculpt v0.1: README copy.md

# Sculpt 🧠

Advanced Multi-Omics Modeling for Elucidating Neuronal Architecture

## Overview

Sculpt is an open source project focused on advanced multi-omics modeling to elucidate neuronal architecture. This project combines cutting-edge techniques in 3D interneuron/neuron image modeling, leveraging work from Google Research and Harvard University.

## Key Features

- Instance segmentation for detailed neuron analysis
- Frame-by-frame image modeling
- 3D neuron prediction from multiomics data
- SEM visualization integration

## Usage

Feel free to use any of the code in this repository. However, we kindly request that you cite this GitHub Project when doing so.

## Methodology

The project utilizes instance segmentation along with frame-by-frame image modeling to create predicted 3D neurons from multiomics data and SEM visualization.

## Collaboration

This project was developed in part through collaboration with Google Research and Harvard University, focusing on 3D interneuron/neuron image modeling.

## Contributing

This is an open source project, and all code and push updates will be posted on this project repository. Contributions are welcome!

## Future Updates

Additional contributors and references will be cited in future updates to this README.


## Contact



---

**Note**: This README is a living document and will be updated as the project evolves.
