## Supplementary material for "A Novel Multi-Omics Deep Learning Framework for Spatiotemporal Cerebral Cortex Localization & Expression": All Code used in building Sculpt v0.1: rnn copy.py

```
import numpy as np
import torch
import torch.nn as nn
import torch.optim as optim
from sklearn.preprocessing import LabelEncoder, MinMaxScaler
from torch.utils.data import DataLoader, TensorDataset

cell_types = ["M4", "M3", "M5", "M1", "M6_t", "M2_t", "M1_t", "M4_t", "M8_t"]
ages = ["Young", "Middle-aged", "Older", "Elderly"]
genes = ["A2M", "ABAT", "ABR", "ACAT1", "ACAT2", "ACO2", "ACOT7", "ACTG1", "ACTN1", "ACTR1A", "ACTR2", "ACTR3"]

data = np.random.rand(len(ages), len(genes))
labels = np.random.randint(0, len(cell_types), size=(len(ages),))

scaler = MinMaxScaler()
data = scaler.fit_transform(data)

label_encoder = LabelEncoder()
labels = label_encoder.fit_transform(labels)

data_tensor = torch.tensor(data, dtype=torch.float32)
labels_tensor = torch.tensor(labels, dtype=torch.long)


class GeneLSTM(nn.Module):
    def __init__(self, input_size, hidden_size, num_classes, num_layers=2, dropout=0.2):
        super(GeneLSTM, self).__init__()
        self.hidden_size = hidden_size
        self.num_layers = num_layers
        self.lstm = nn.LSTM(input_size, hidden_size, num_layers, batch_first=True, dropout=dropout, bidirectional=True)
        self.fc = nn.Linear(hidden_size * 2, num_classes)

    def forward(self, x):
        h0 = torch.zeros(self.num_layers * 2, x.size(0), self.hidden_size)
        c0 = torch.zeros(self.num_layers * 2, x.size(0), self.hidden_size)
        out, _ = self.lstm(x, (h0, c0))
        out = self.fc(out[:, -1, :])
        return out


input_size = len(genes)
hidden_size = 64
num_classes = len(cell_types)
num_layers = 3
model = GeneLSTM(input_size, hidden_size, num_classes, num_layers)

criterion = nn.CrossEntropyLoss()
optimizer = optim.Adam(model.parameters(), lr=0.001)

epochs = 150
batch_size = 8
dataset = TensorDataset(data_tensor, labels_tensor)
dataloader = DataLoader(dataset, batch_size=batch_size, shuffle=True)

scheduler = optim.lr_scheduler.StepLR(optimizer, step_size=50, gamma=0.5)

best_loss = float('inf')
early_stopping_patience = 20
patience_counter = 0

for epoch in range(epochs):
    model.train()
    running_loss = 0.0
    for i, (inputs, targets) in enumerate(dataloader):
        optimizer.zero_grad()
        outputs = model(inputs.unsqueeze(1))
        loss = criterion(outputs, targets)
        loss.backward()
        optimizer.step()
        running_loss += loss.item()

    scheduler.step()

    avg_loss = running_loss / len(dataloader)

    if avg_loss < best_loss:
        best_loss = avg_loss
        patience_counter = 0
    else:
        patience_counter += 1

    if patience_counter >= early_stopping_patience:
        print("Early stopping triggered")
        break

    if epoch % 10 == 0:
        print(f"Epoch [{epoch}/{epochs}], Loss: {avg_loss:.4f}")

print("Training complete.")
```
