## Supplementary material for "A Novel Multi-Omics Deep Learning Framework for Spatiotemporal Cerebral Cortex Localization & Expression": All Code used in building Sculpt v0.1: smpcnn copy.py

```
import os
import torch
import cv2
import numpy as np
import segmentation_models_pytorch as smp
from torch.utils.data import Dataset, DataLoader
from torch import nn
import torch.optim as optim


# Define Dataset
class NeuronalDataset(Dataset):
    def __init__(self, root_dir, transform=None):
        self.root_dir = root_dir
        self.transform = transform
        self.data = []
        self.class_names = sorted(os.listdir(root_dir))
        for label, class_dir in enumerate(self.class_names):
            class_path = os.path.join(root_dir, class_dir)
            for file in os.listdir(class_path):
                img_path = os.path.join(class_path, file)
                self.data.append((img_path, label))

    def __len__(self):
        return len(self.data)

    def __getitem__(self, idx):
        img_path, label = self.data[idx]
        img = cv2.imread(img_path, cv2.IMREAD_GRAYSCALE)
        img = cv2.resize(img, (128, 128))
        img = np.expand_dims(img, axis=0)  # (1, 128, 128)
        img = torch.tensor(img, dtype=torch.float32) / 255.0

        # Create a segmentation mask (you may need to adjust this depending on your data format)
        label_mask = np.zeros((128, 128), dtype=np.int64)  # Ensure labels are in the right shape
        label_mask[:] = label  # Set the entire mask to the class index

        label_mask = torch.tensor(label_mask, dtype=torch.long)

        return img, label_mask


# Load Dataset
dataset = NeuronalDataset(root_dir="/Users/ganeshtalluri/PycharmProjects/Sculpt/Patches")
dataloader = DataLoader(dataset, batch_size=4, shuffle=True)

# Define Model
model = smp.Unet(encoder_name="resnet34", in_channels=1, classes=5)
device = torch.device("cuda" if torch.cuda.is_available() else "cpu")
model.to(device)

# Define Loss and Optimizer
criterion = nn.CrossEntropyLoss()
optimizer = optim.Adam(model.parameters(), lr=0.001)

# Training Loop
num_epochs = 10
for epoch in range(num_epochs):
    total_loss = 0
    correct_pixels = 0
    total_pixels = 0
    for images, labels in dataloader:
        images, labels = images.to(device), labels.to(device)
        optimizer.zero_grad()
        outputs = model(images)
        loss = criterion(outputs, labels)  # Labels are now in the correct shape
        loss.backward()
        optimizer.step()
        total_loss += loss.item()

        # Calculate pixel-wise accuracy
        _, predicted = torch.max(outputs, 1)  # Get the predicted class for each pixel
        correct_pixels += (predicted == labels).sum().item()  # Count correct pixels
        total_pixels += labels.numel()  # Total number of pixels in the batch

    accuracy = 100 * correct_pixels / total_pixels  # Calculate pixel-wise accuracy
    print(f"Epoch [{epoch + 1}/{num_epochs}], Loss: {total_loss / len(dataloader):.4f}, Accuracy: {accuracy:.2f}%")

# Save Model
torch.save(model.state_dict(), "smp_unet_model.pth")
```
