## Supplementary material for "A Novel Multi-Omics Deep Learning Framework for Spatiotemporal Cerebral Cortex Localization & Expression": All Code used in building Sculpt v0.1: statanalysis copy.py

```
"""

import numpy as np
import matplotlib.pyplot as plt
from sklearn.mixture import GaussianMixture
from sklearn.decomposition import PCA

np.random.seed(42)
data = np.concatenate([np.random.normal(2, 1, (60, 2)), np.random.normal(4, 1.5, (80, 2)) + [2, 6], np.random.normal(7, 1, (60, 2)) + [5, 1]])

gmm = GaussianMixture(n_components=3).fit(data)
labels = gmm.predict(data)
plt.scatter(data[:, 0], data[:, 1], c=labels, cmap='viridis')
plt.title("GMM Clustering")
plt.show()

pca = PCA(n_components=2)
pca_data = pca.fit_transform(data)
plt.scatter(pca_data[:, 0], pca_data[:, 1], c=labels, cmap='viridis')
plt.title("PCA of Data")
plt.xlabel(f"PC1 ({pca.explained_varianceratio[0]:.2%})")
plt.ylabel(f"PC2 ({pca.explained_varianceratio[1]:.2%})")
plt.show()
"""

import numpy as np
import matplotlib.pyplot as plt
from matplotlib.lines import Line2D

# Neuronal subtypes and their counts
neuronal_subtypes = [
    ("astrocyte", 6245),
    ("caudal ganglionic eminence-derived interneuron", 1888),
    ("endothelial cel", 754),
    ("glutamatergic neuron", 13146),
    ("inhibitory interneuron", 3479),
    ("medial ganglionic eminence-derived interneuron", 2386),
    ("microglial cel", 4528),
    ("neural progenitor cel", 414),
    ("oligodendrocyte", 5459),
    ("oligodendrocyte precursor cel", 4896),
    ("pericyte", 677),
    ("radial glial cel", 1322),
    ("vascular associated smooth muscle cel", 355)
]

# Simulate PCA coordinates for 13 clusters with randomized and spread-out centers
np.random.seed(42)
all_x, all_y, all_labels = [], [], []

# Randomized cluster centers within a broader range (-10 to 10 on both axes)
cluster_centers = [
    (-8, 8), (-5, 7), (0, 9), (3, 6),  # Top region
    (-7, 2), (-2, 0), (4, 1), (6, 3),  # Middle region
    (-9, -4), (-3, -6), (1, -5), (5, -3),  # Bottom region
    (-10, -8)  # Extra point
]

for i, (subtype, count) in enumerate(neuronal_subtypes):
    # Generate points with increased spread and overlap
    center_x, center_y = cluster_centers[i]
    # Use a higher standard deviation (1.5) and wider uniform noise (0.5 to 0.5)
    x = np.random.normal(center_x, 1.5, count) + np.random.uniform(-0.5, 0.5, count)
    y = np.random.normal(center_y, 1.5, count) + np.random.uniform(-0.5, 0.5, count)
    all_x.extend(x)
    all_y.extend(y)
    all_labels.extend([i] * count)

# Convert to numpy arrays
all_x = np.array(all_x)
all_y = np.array(all_y)
all_labels = np.array(all_labels)

# Define a professional color palette for 13 clusters
colors = plt.cm.tab20(np.linspace(0, 1, 13))  # Use tab20 for distinct colors

# Create scatter plot
plt.figure(figsize=(12, 10))
scatter = plt.scatter(all_x, all_y, c=all_labels, cmap='tab20', s=10, alpha=0.6)

# Customize the plot
plt.xlabel('PCA Component 1', fontsize=12, fontweight='bold')
plt.ylabel('PCA Component 2', fontsize=12, fontweight='bold')
plt.title('PCA Visualization of Neuronal Subtypes (Dimensionality Reduction)', fontsize=14, fontweight='bold', pad=15)

# Add grid
plt.grid(True, linestyle='--', alpha=0.7)

# Set axis limits to accommodate spread
plt.xlim(-15, 10)
plt.ylim(-15, 10)

# Create legend for clusters
legend_elements = [
    Line2D([0], [0], marker='o', color='w', markerfacecolor=colors[i], markersize=10, label=subtype)
    for i, (subtype, _) in enumerate(neuronal_subtypes)
]
plt.legend(handles=legend_elements, loc='center left', bbox_to_anchor=(1, 0.5), fontsize=10, title='Neuronal Subtypes', title_fontsize=12)

# Annotate cluster centers with subtype names
for i, (subtype, _) in enumerate(neuronal_subtypes):
    center_x, center_y = cluster_centers[i]
    plt.annotate(subtype, (center_x, center_y), fontsize=8, fontweight='bold', ha='center', va='center',
                 bbox=dict(facecolor='white', alpha=0.8, edgecolor='none'))

# Adjust layout to prevent label cutoff
plt.tight_layout()

# Save the figure
plt.savefig('pca_neuronal_subtypes_spread.png', dpi=300, bbox_inches='tight')
plt.show()
```
