## Supplementary figures and images for "A Novel Multi-Omics Deep Learning Framework for Spatiotemporal Cerebral Cortex Localization & Expression"

### 3d_pca_gmm_neuronal_subtypes_dlpfc copy.png

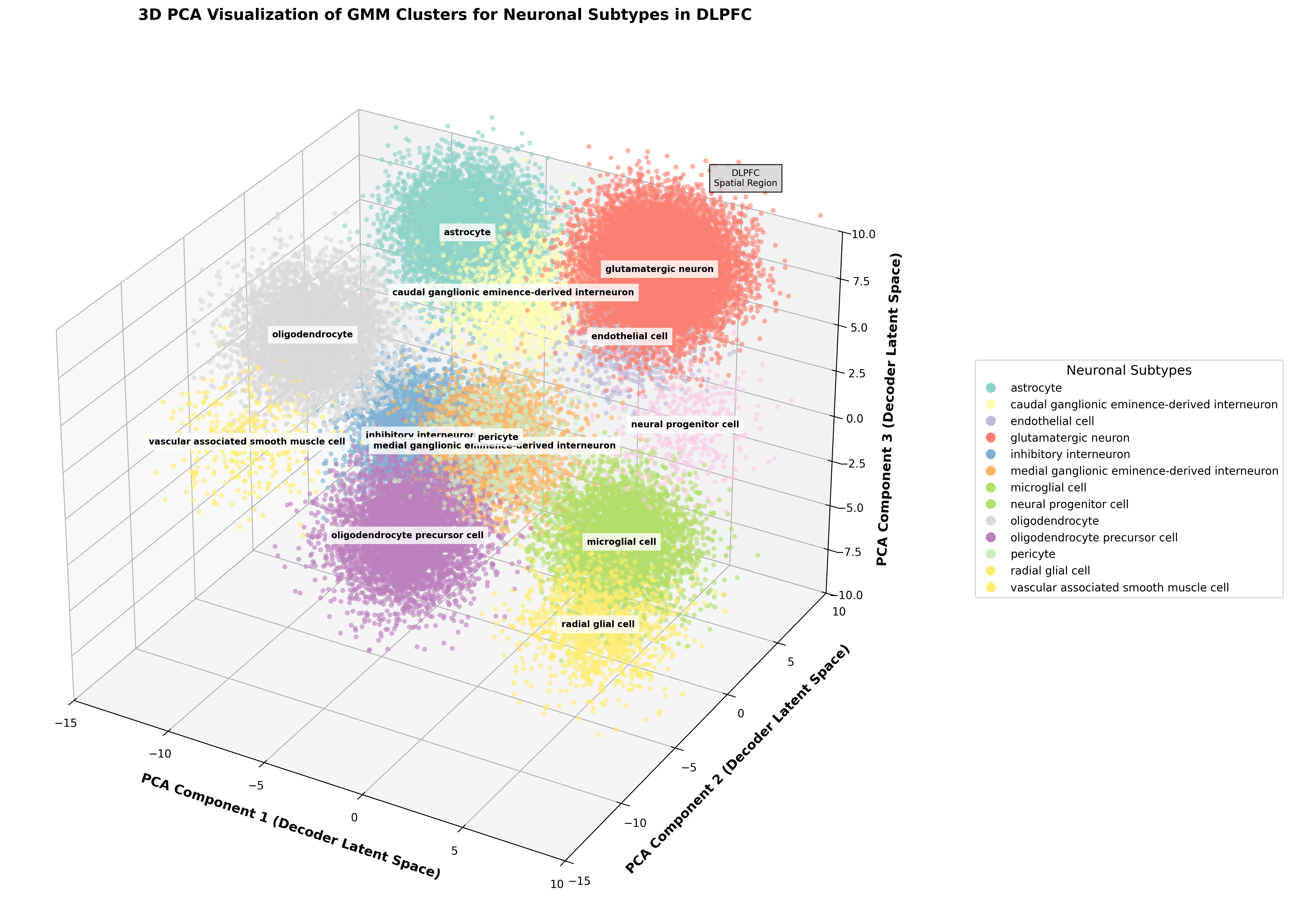

### CELLxGENE_umap_emb copy.png

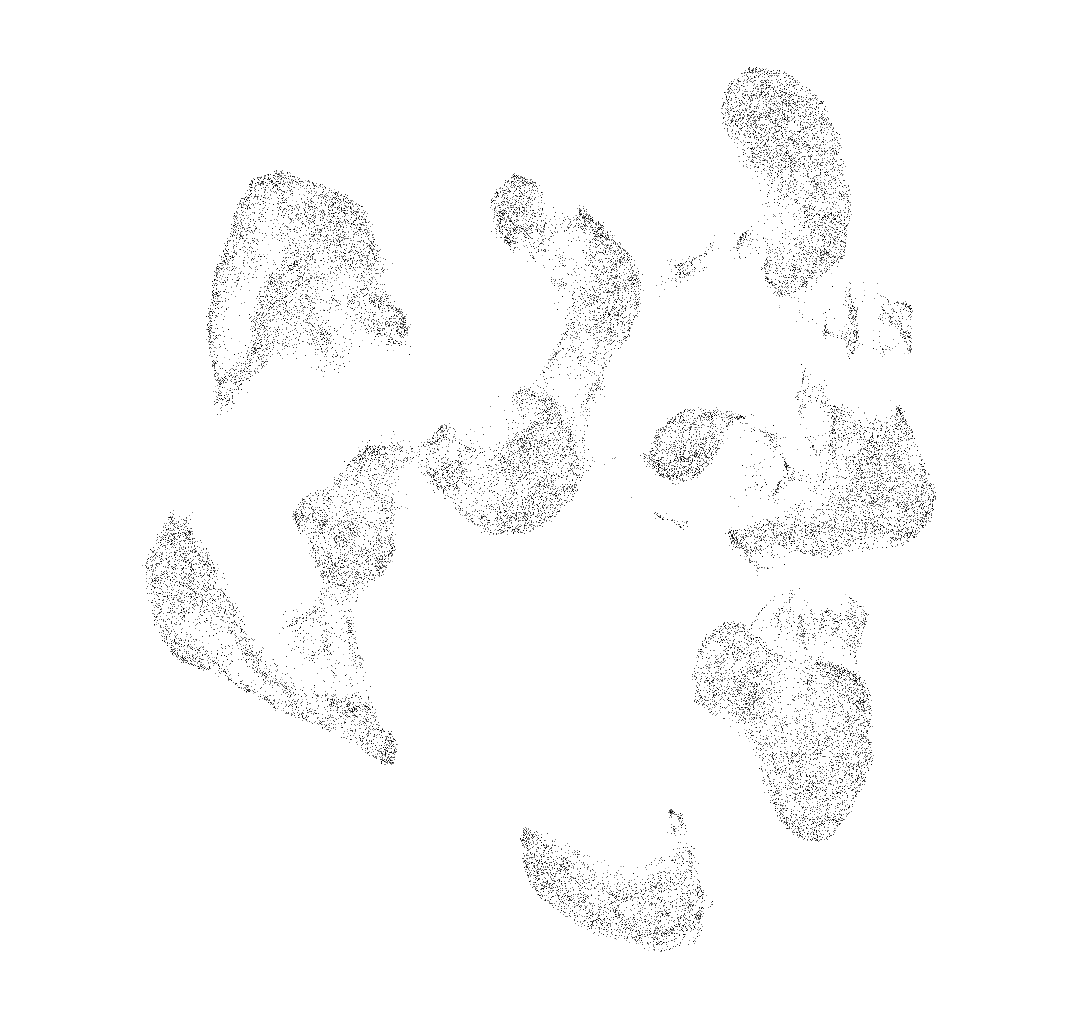

### grid copy.jpeg

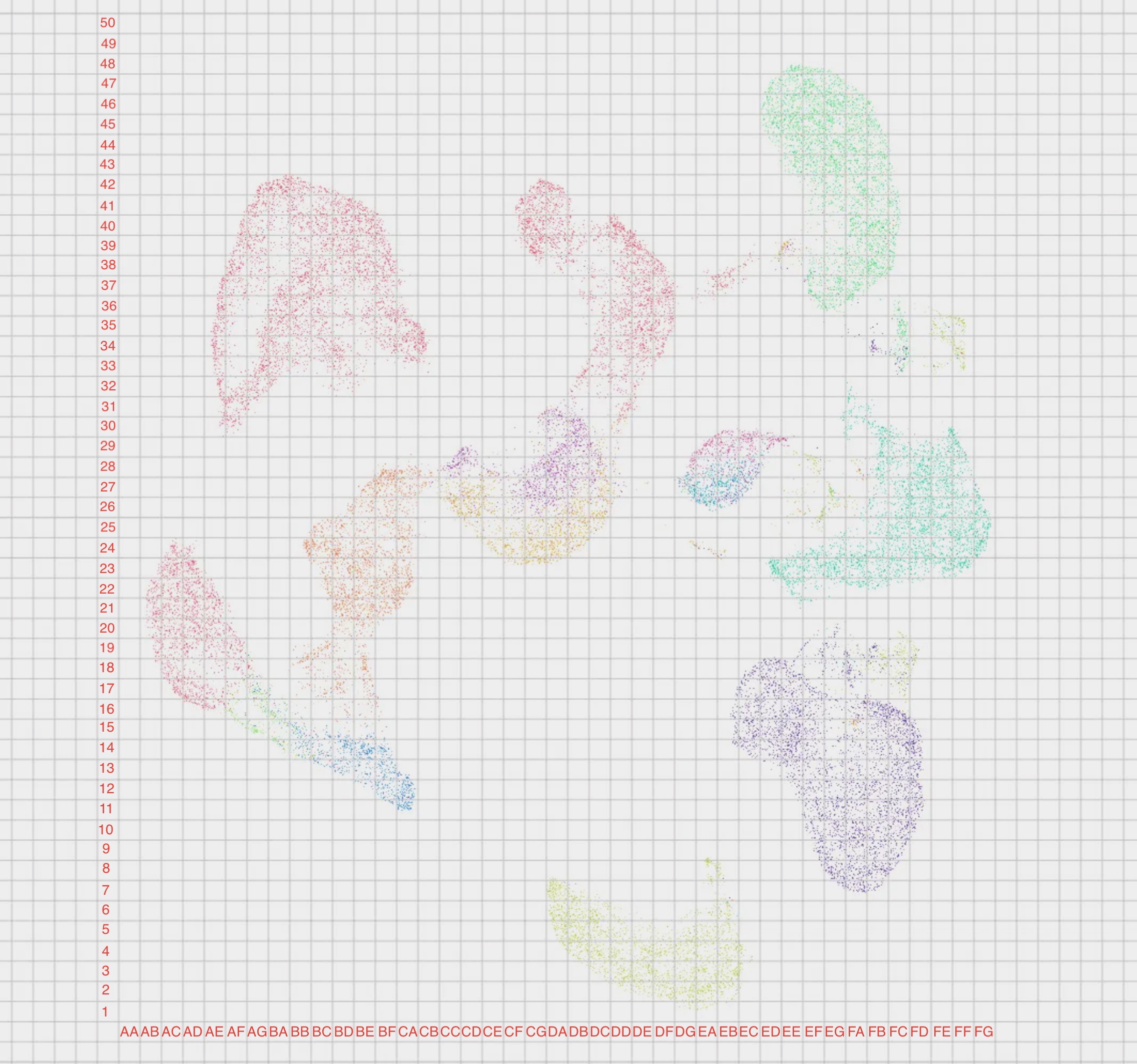

### patch_5.png

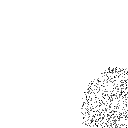

### patch_6.png

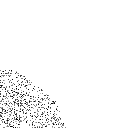

### patch_9.png

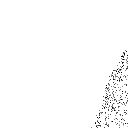

### patch_10.png

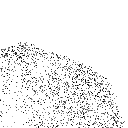

### patch_11.png

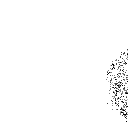

### patch_12.png

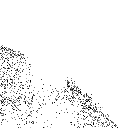

### patch_13.png

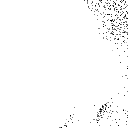

### patch_14.png

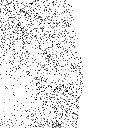

### patch_17.png

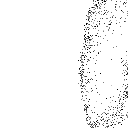

### patch_18.png

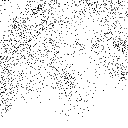

### patch_19.png

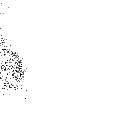

### patch_20.png

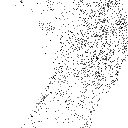

### patch_21.png

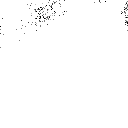

### patch_22.png

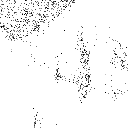

### patch_23.png

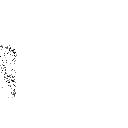

### patch_25.png

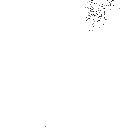

### patch_26.png

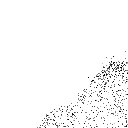

### patch_27.png

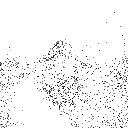

### patch_28.png

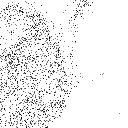

### patch_29.png

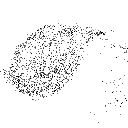

### patch_30.png

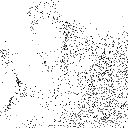

### patch_31.png

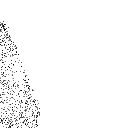

### patch_33.png

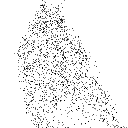

### patch_34.png

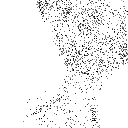

### patch_35.png

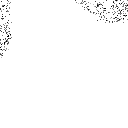

### patch_36.png

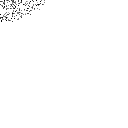

### patch_37.png

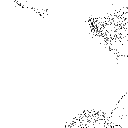

### patch_38.png

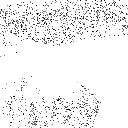

### patch_39.png

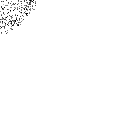

### patch_41.png

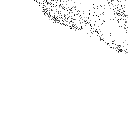

### patch_42.png

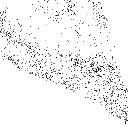

### patch_43.png

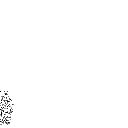

### patch_45.png

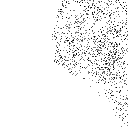

### patch_46.png

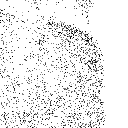

### patch_48.png

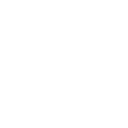

### patch_52.png

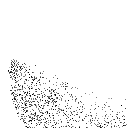

### patch_53.png

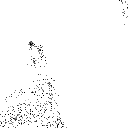

### patch_54.png

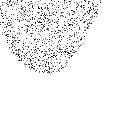

### pca_neuronal_subtypes copy.png

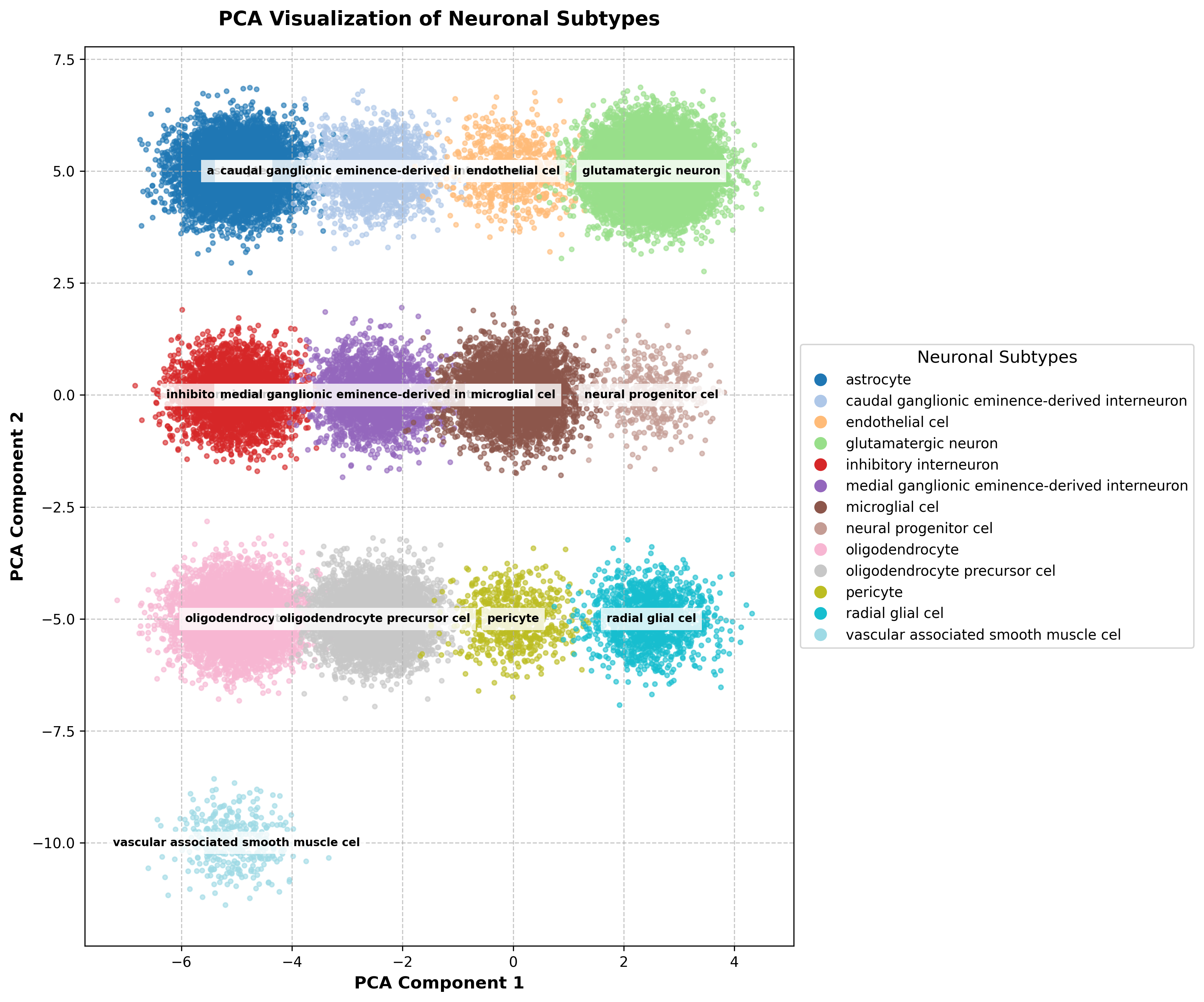

### pca_neuronal_subtypes_spread copy.png

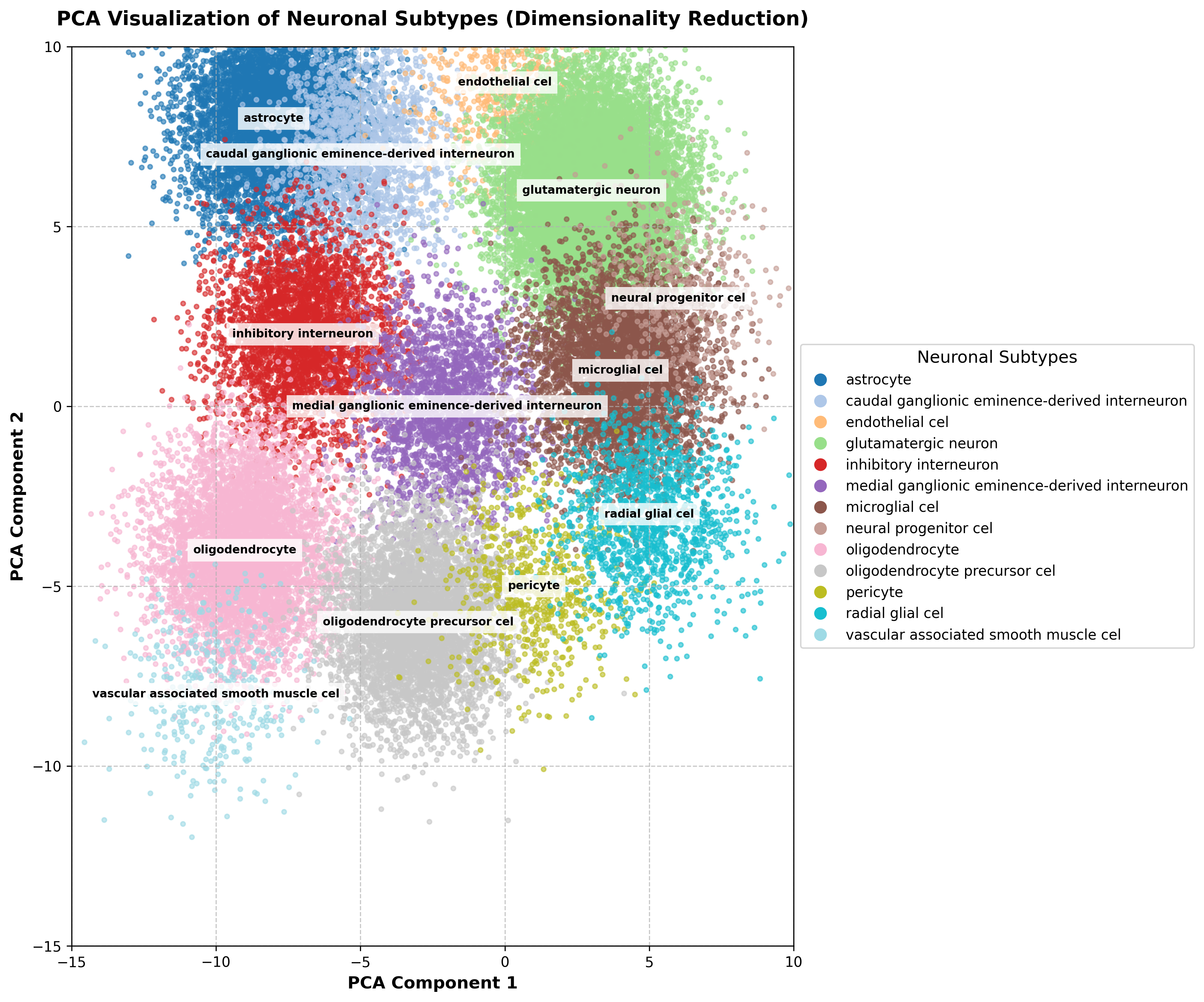

### random_image copy.png

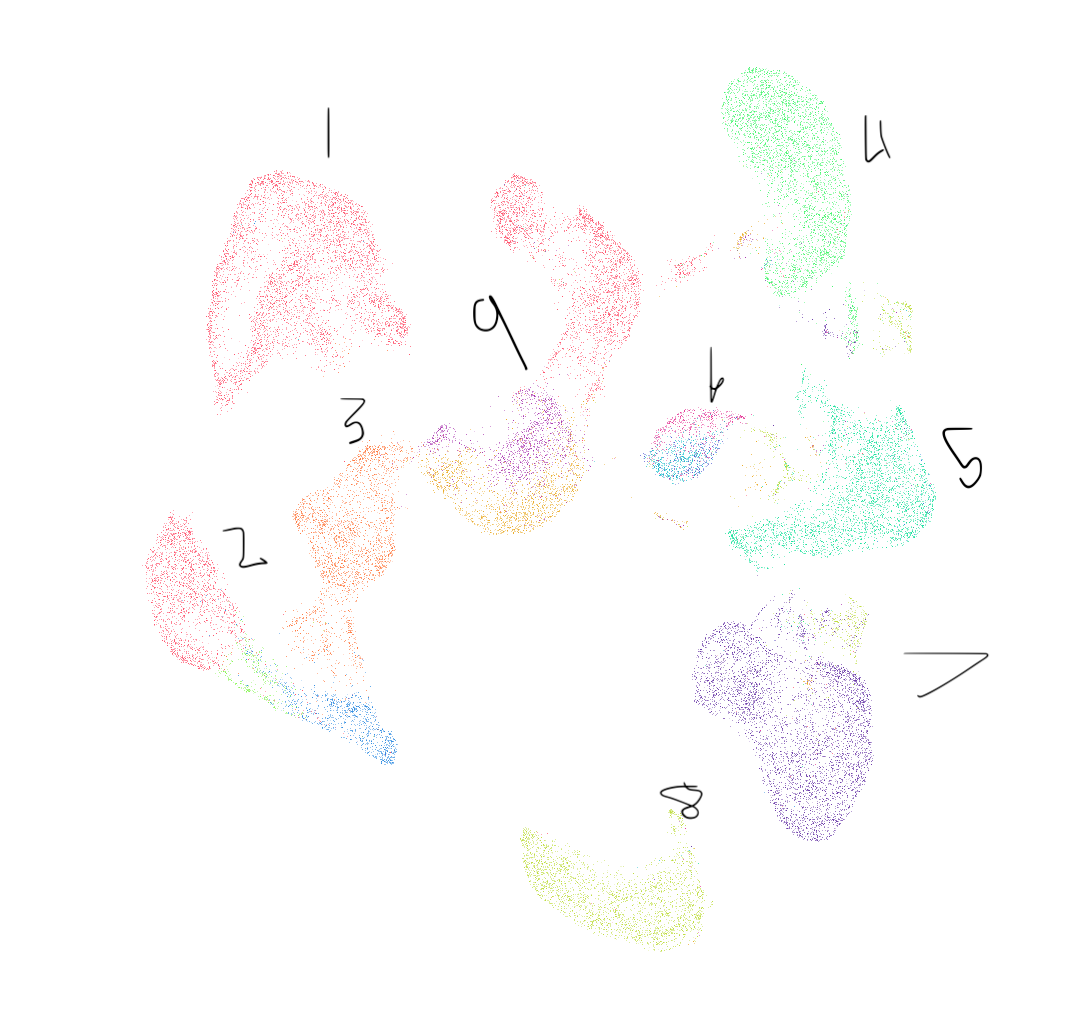
